# Automatic ICD coding using LLMs: a systematic review

**DOI:** 10.1101/2025.07.30.25330916

**Authors:** Adi Gershon, Shelly Soffer, Girish N Nadkarni, Eyal Klang

## Abstract

**Background:** Manual assignment of International Classification of Diseases (ICD) codes is error-prone. Transformer-based large language models (LLMs) have been proposed to automate coding, but their accuracy and generalizability remain uncertain.

**Methods:** We performed a systematic review registered with PROSPERO (CRD42024576236) and reported according to PRISMA guidelines. PubMed, Embase, and Google Scholar were searched through January 2025 for peer-reviewed studies that evaluated an LLM (e.g., BERT, GPT) for ICD coding and reported at least one performance metric. Two reviewers independently screened articles, extracted data, and assessed methodological quality with the Joanna Briggs Institute Critical Appraisal Checklist for Analytical Cross-Sectional Studies. Outcomes included micro-F1, macro-F1, accuracy, precision, recall, and AUC, capturing both overall predictive performance and sensitivity to rare ICD codes.

**Results:** Of 590 records screened, 35 studies met the inclusion criteria. 24 assessed general-purpose coding across broad clinical text, 10 focused on specific clinical contexts, and 11 addressed multilingual interoperability; some studies belonged to more than one theme. Median micro-F1 for frequent codes was 0.79 (range, 0.73–0.94), exceeding that of legacy machine-learning baselines in all comparative studies. Performance for infrequent codes was lower (median macro-F1, 0.42) but improved modestly with data augmentation, contrastive retrieval, or graph-based decoders. Only 1 study used federated learning across institutions, and 3 conducted external validation. The risk-of-bias assessment rated 18 studies (51%) as moderate, primarily due to unclear blinding of assessors and selective reporting.

**Conclusions:** LLM-based systems reliably automate common ICD codes and frequently match or surpass professional coders, but accuracy declines for rare diagnoses, and external validation is scant. Prospective, multicenter trials and transparent reporting of prompts and post-processing rules are required before clinical deployment.

## INTRODUCTION

International Classification of Diseases (ICD) codes play an important role in healthcare systems. They drive billing processes, inform clinical decision-making, and facilitate health policy planning. Traditionally, ICD coding relied on manual data entry, a process that can be time-consuming and prone to error. As patient records grow in volume and complexity, there is a growing need for efficient automated solutions.

In recent years, artificial intelligence (AI) methods have shown promise in tackling complex tasks within healthcare informatics, including ICD coding automation. Early machine learning approaches, such as rule-based algorithms, random forests, and support vector machines, demonstrated the potential to enhance coding speed and consistency. However, these models often struggled to capture nuanced clinical language and contextual information.

Large Language Models (LLMs), such as Bidirectional Encoder Representations from Transformers (BERT), and generative pre-trained transformer (GPT), have emerged as powerful tools in natural language processing (NLP). Their ability to parse and interpret free-text clinical documentation suggests that LLM-based approaches could improve ICD coding performance (**Figure 1**).

**Figure 1.**
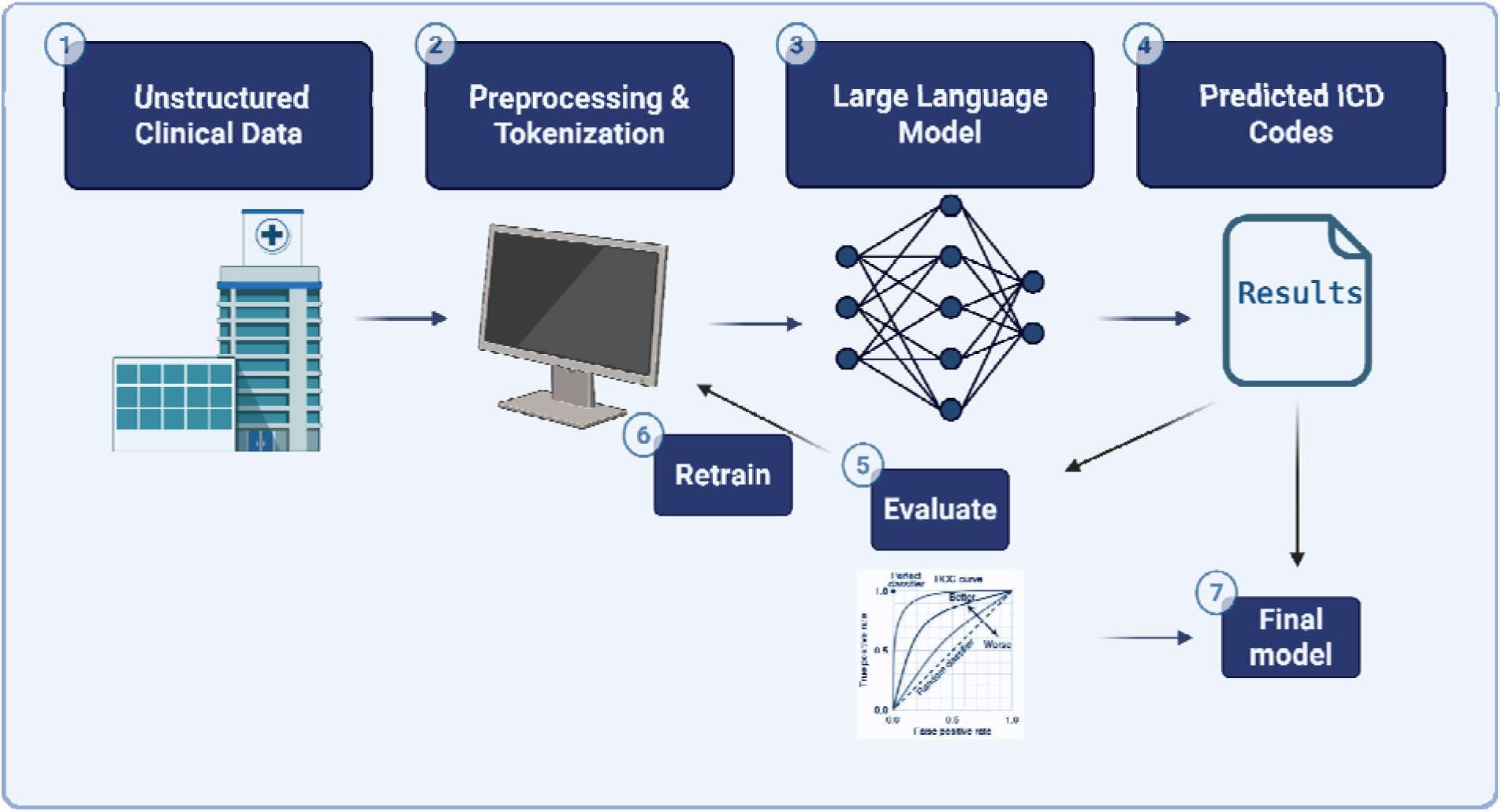
Schematic Workflow of Automated ICD Coding Using Machine Learning. Illustration of the pipeline for automated ICD coding using LLMs: 1. unstructured clinical text (EHR notes, discharge summaries). 2. Preprocessing and tokenization of the data. 3. Fine-tuning a large language model on labeled examples. 4. Generation of predicted ICD codes. 5. performance assessment against gold standards using accuracy, precision, recall and F1. 6. iterative error analysis and model refinement; 7. Final Model. validated LLM ready for deployment.

**Figure 2.**
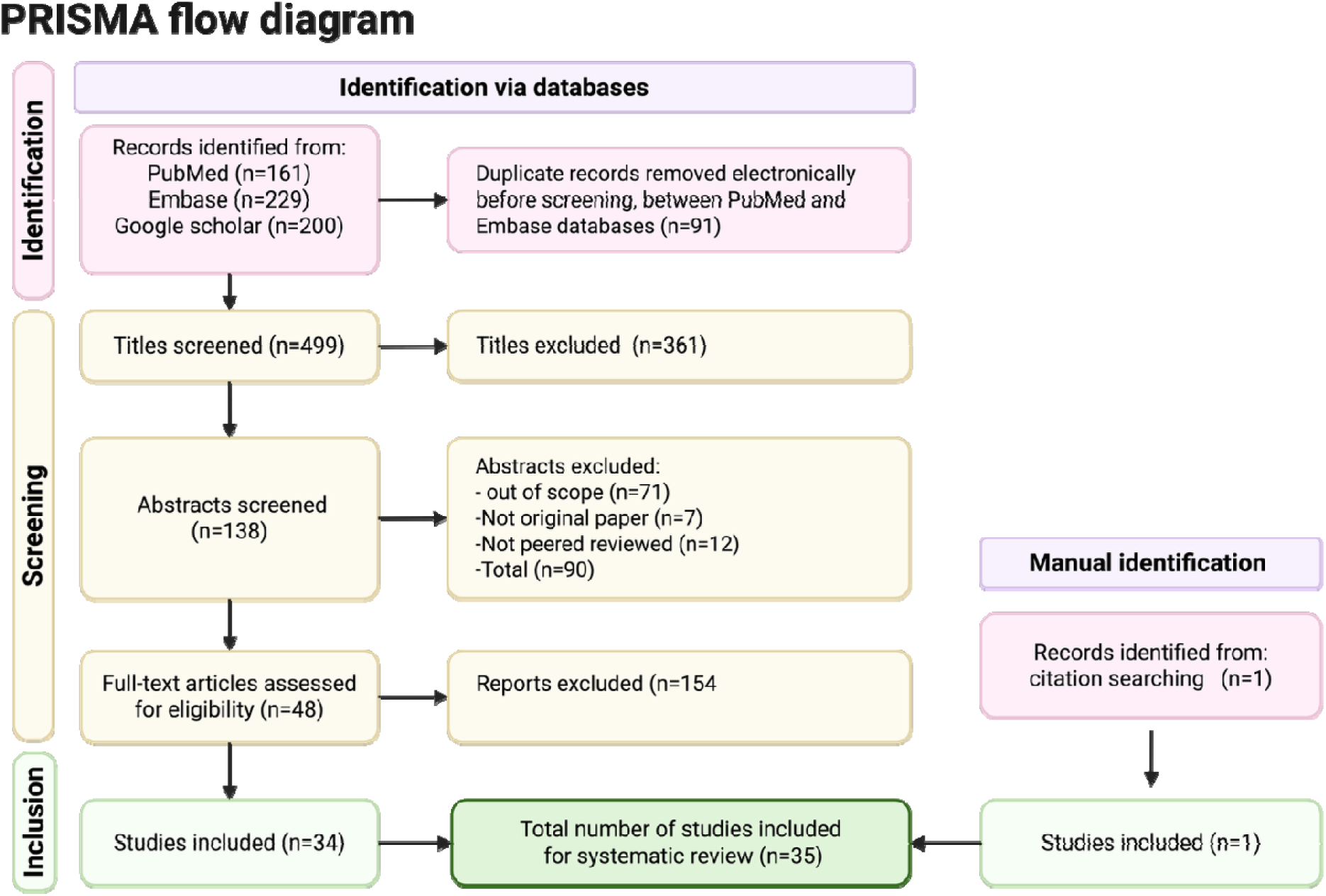
PRISMA flow diagram of study selection. 590 records were identified (PubMed 161; Embase 229; Google Scholar 200). After removing 91 duplicates, 499 titles were screened (361 excluded), 138 abstracts were assessed (90 excluded), and 48 full texts were reviewed (14 excluded). A total of 34 studies were included, plus 1 from citation searching (35 total).

We aimed to synthesize published evidence on the accuracy of LLMs for automatic ICD coding, compare their performance with manual and legacy automated methods, and identify limitations that should direct future model development and validation.

## METHODS

This systematic review was conducted by the Preferred Reporting Items for Systematic Reviews (PRISMA) guidelines. The review protocol was registered prospectively with PROSPERO (CRD42024576236).

### Search Strategy

A systematic literature search was conducted to identify peer-reviewed studies evaluating the use of Large Language Models (LLMs) for automatic ICD coding. The search was performed in PubMed, Embase and Google Scholar databases in January 2025. Search terms included combinations of keywords such as “ICD coding”, “large language models”, “GPT”, “BERT”, “transformer models”, “ChatGPT”, and “medical coding” (**Table S1**). Articles published in English were included. Additional studies were identified by manually screening the reference lists of included articles.

### Inclusion and Exclusion Criteria

Studies were included if they: 1) Evaluated the application of a Large Language Model (e.g., BERT, GPT, Google Gemini, Microsoft Bing) for ICD coding, 2) Were original research, peer-reviewed, and published in English, 3) Reported at least one performance metric (accuracy, F1 score, precision, recall) 4) Used clinical or administrative datasets involving ICD classification.

Studies were excluded if they: 1) Focused on manual coding without the use of LLMs, 2) Used other coding systems (e.g., CPT, SNOMED) without reference to ICD, 3) Were reviews, editorials, conference abstracts, or non-peer-reviewed sources. (**Table S2**)

To capture the practical application of LLMs in ICD coding, we extracted representative examples of prompt–response pairs from included studies. (**Table S3**) These examples illustrate how models interpret clinical descriptions to generate structured ICD-10 codes.

### Study Selection

Two reviewers independently screened titles and abstracts. Studies that appeared to meet the inclusion criteria, or those where eligibility was unclear, were reviewed in full text. Discrepancies were resolved through discussion or consultation with a third reviewer.

### Data Extraction

A standardized data extraction form was used to collect key variables from each included study. Extracted data included: author, year, model architecture (e.g., GPT, BERT), type of LLM used, data source, data size, preprocessing techniques, fine-tuning, comparator methods (if any), evaluation metrics, and key outcomes. Data were extracted independently by two reviewers, with disagreements resolved through discussion.

### Risk of bias and quality assessment

We evaluated risk of bias using the Joanna Briggs Institute (JBI) Critical Appraisal Checklist for Analytical Cross-Sectional Studies; two reviewers independently rated each study across key domains (sample selection, measurement validity, confounding) and resolved any discrepancies by consensus.

## Results

The database search retrieved 590 records. After title/abstract screening and fulll7ltext review 35 studies met all inclusion criteria. We grouped these studies into three thematic categories (**Figure 3**). Below, we summarize the findings in each category, with overlapping studies discussed in each relevant section.

**Figure 3.**
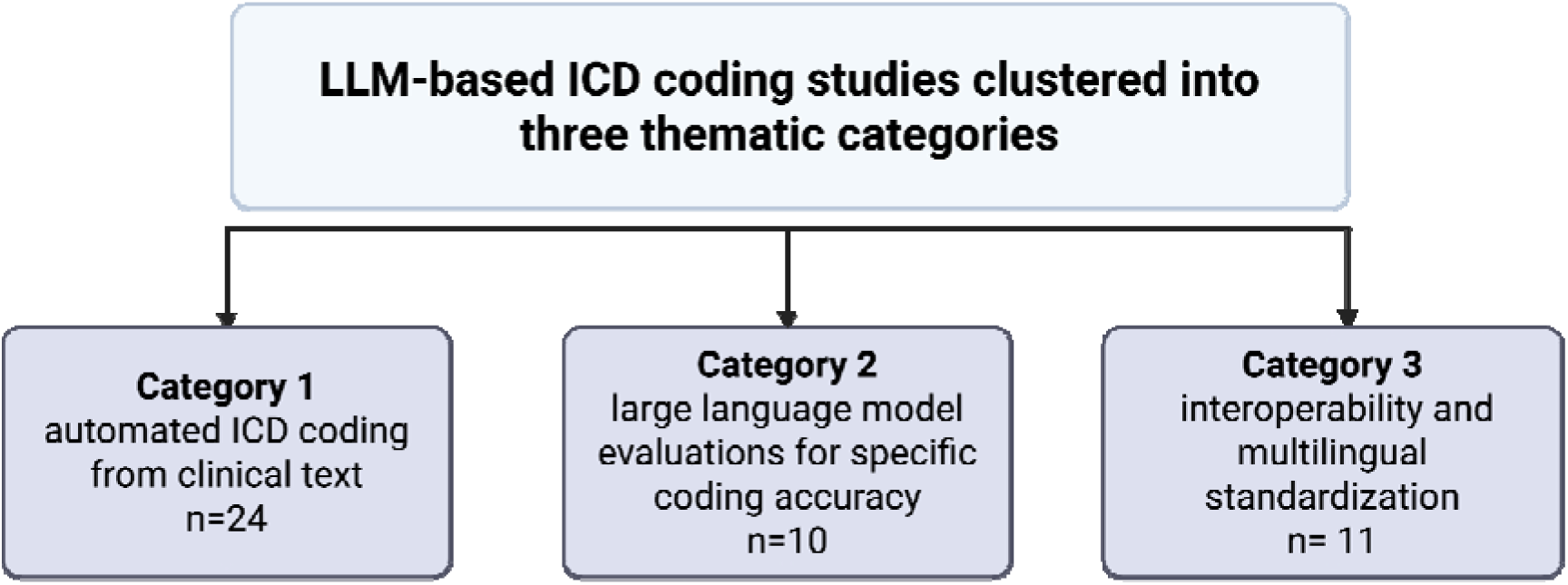
Thematic clustering of included LLM-based ICD coding studies. Among 590 records identified, 35 met inclusion criteria and were assigned to three overlapping themes. Category 1 (n = 24): automated ICD coding from broad clinical text. Category 2 (n = 10): LLM evaluations in specific clinical contexts. Category 3 (n = 11): interoperability and multilingual standardization.

### Automated ICD Coding

The first category includes 24 studies that evaluate general-purpose ICD-10 coding using transformer-based models applied to broad clinical datasets. These studies aimed to maximize coverage across common and rare ICD codes, often using discharge summaries or multi-section EHR notes. Fine-tuned transformer encoders such as BioBERT, ClinicalBERT, and XLNet consistently achieved high performance for frequent codes, with micro-F1 scores typically ranging from 0.73 to 0.83 ^1–7^ . When long-sequence models like ClinicalLongformer were used to capture full discharge narratives, accuracy improved to 94%, especially in ICU datasets ^8^. However, when rare ICD codes were included, macro-level performance dropped substantially, underscoring the ongoing challenge of coding low-frequency labels ^1^. To address this, several complementary strategies were used. Candidate-reduction methods like contrastive retrieval and reranking improved top-5 prediction accuracy to 0.835 ^9^. Rule-based filtering of implausible code combinations boosted BioBERT’s F1 by three points ^10^ .Synthetic data generation using GPT-3.5 nearly doubled macro-F1 for rare codes through targeted augmentation ^11^. Federated learning enabled model training across hospitals without data sharing, preserving privacy while maintaining strong micro-F1 scores ^12^.Graph-aware decoders such as transformer-GCN hybrids and probabilistic label trees raised micro-F1 on MIMIC benchmarks by explicitly modeling code dependencies ^13,14^, while an enhanced cosine-similarity improved micro-F1 to a PLM-ICD baseline^15^. Real-world evaluations underline practical value: a GPT-2–assisted coder deployed in Taiwan achieved κ ≈ 0.71 with professional coders, a GPT- 4-enhanced hierarchical framework sustained F1 ≈ 0.89, and fine-tuning GPT-3.5 on only 1,000 notes more than doubled accuracy on common codes ^16–18^

### Specific ICD Coding

The next category focuses on evaluations of large language models (LLMs) for ICD-10 coding in specific clinical contexts. Examples include gastrointestinal discharge summaries, death certificates, nephrology case reports, and retina clinic vignettes. Most studies in this group applied LLMs to focused medical tasks, often in one language, to test how well these models perform in realistic domain settings.

Across studies, performance was high when models were applied to structured, in-domain data. Macro F1 scores generally ranged from 0.70 to 0.96, and top-5 predictions were used to support coders by narrowing code options^19–22^ In simulated scenarios, accuracy reached as high as 99 percent ^23^. In Zambetta et al, hybrid system outperformed GPT-3.5 on a mortality coding task ^24^.

Class imbalance remains a major bottleneck across studies—rare codes continue to be under- predicted, and although methods such as influence-balanced loss and per-label attention deliver modest F1 gains, absolute full-code F1 scores for infrequent labels remain low one paper report an increase from 34.1% to 35.0% F1 ^22^. Blanco et al. observe only small Macro-F1 boosts into the 40–50% range ^25^. Some models circumvented this issue by limiting predictions to frequent codes ^21^.

Another common finding was the value of domain- and language-specific models: pretraining or fine-tuning on in-domain clinical text consistently improved coding performance ^19,20,25,26^.

Swedish BERT fine-tuned on 56 M tokens of discharge summaries raised “strict” full-code F1 from 0.43 to 0.94 ^20^.

Finally, several studies showed Prompt-based LLMs such as GPT-4, even without any fine-tuning, have matched or outperformed traditional baselines, whether human reviewers, rule-based coding algorithms, or earlier LLM versions on narrow, well-defined tasks when driven by carefully designed prompts. This suggests LLMs can be effective in real-world use cases with minimal setup, though reproducibility depends on transparent prompts and access conditions^23,27,28^.

### Interoperability and Multilingual Standardization

The third category includes studies that address multilingual ICD-10 coding and interoperability across different healthcare systems and languages. These models aim to generalize across diverse datasets, languages, and documentation formats. A consistent strength across studies was the successful adaptation of LLMs to non-English languages, including Chinese, Spanish, Portuguese, Czech, Swedish, and French ^19,22,25,29–32^. Recent bilingual work further extends coverage to Thai clinical narratives ^33,34^. Some models achieved high performance at the chapter or block level, with macro-F1 ranging from 0.72 to 0.95 ^19,22^. Several studies emphasized in-domain pretraining and language-specific tuning as critical for performance, particularly in lower-resource languages like Czech and Swedish ^22,29^. However, full-code prediction remained challenging, with F1 scores dropping below 40 percent in detailed ICD categories despite sophisticated architectures and class balancing ^22,31^ . A shared limitation was the difficulty of handling rare codes and long sequences, which affected generalization across datasets ^25,34^. Models like PlaBERT improved multilingual accuracy using per-label attention but still required careful alignment between languages and codes ^25^.Large-scale national implementations and hybrid coding pipelines further demonstrated real-world feasibility, as shown by French mortality surveillance ^24^. Task-tailored architectures, such as summarization-based coding for oncology discharge summaries, likewise sustained strong micro-AUC despite narrow code ranges ^26^. Together, these findings highlight that multilingual ICD coding is achievable, but success depends on combining pretraining on local clinical text with targeted model adaptation and careful handling of rare or complex labels.

## DISCUSSION

Transformer-based LLMs outperform legacy automated systems and, in selected settings, match or exceed manual coders. Micro-F1 values clustered near 0.75 for high-frequency codes^1,5,6^.

Important gaps remain. Performance fell sharply for low-prevalence codes despite data augmentation, contrastive retrieval, and class-balanced loss ^22,24,25^. Most studies relied on retrospective datasets from single hospitals ^2,4–10,13–18,27,29,30^, only one used federated learning across site ^12(p1)^. External validation, workflow studies, and cost analyses were rare ^27,35^. Model transparency was uneven: prompts, temperature settings, and post-processing rules were often missing, limiting reproducibility ^35^.

Clinical deployment therefore demands caution. Coders remain accountable for final assignment and must understand model boundaries. Rare diagnoses, crosswalk errors, and jurisdiction-specific billing rules still require expert review. Regulators will expect evidence that automated coding does not amplify disparities in reimbursement or quality metrics.

This review has limitations. We searched only PubMed, Embase and Google scholar and limited inclusion to English-language articles indexed through January 2025 relevant non-indexed or non-English studies may have been missed. Heterogeneity in datasets, code granularity, and outcome metrics precluded meta-analysis, so performance estimates are descriptive. Finally, several reports lacked sufficient methodological detail for full appraisal.

Future work should focus on four areas: prospective trials comparing coder-only, model-only, and hybrid workflows; benchmark sets with adjudicated rare codes; mandatory open reporting of prompts and post-processors; and privacy-preserving multicenter learning to improve coverage of uncommon conditions.

In conclusion, LLMs have advanced automatic ICD coding from proof-of-concept to near- production performance for common codes, but rigorous external validation and safeguards for rare and complex diagnoses remain essential.

**Table 1:**
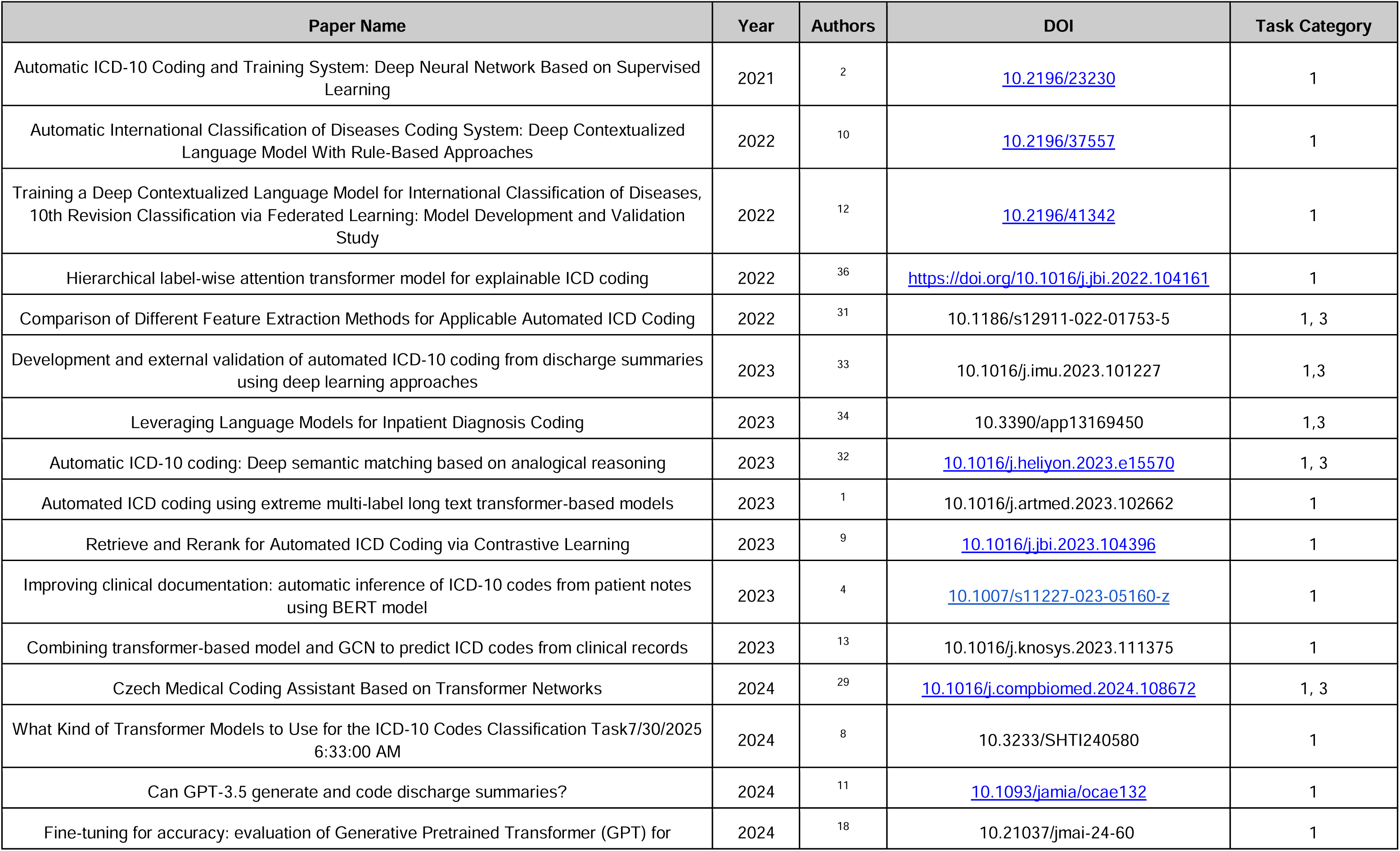
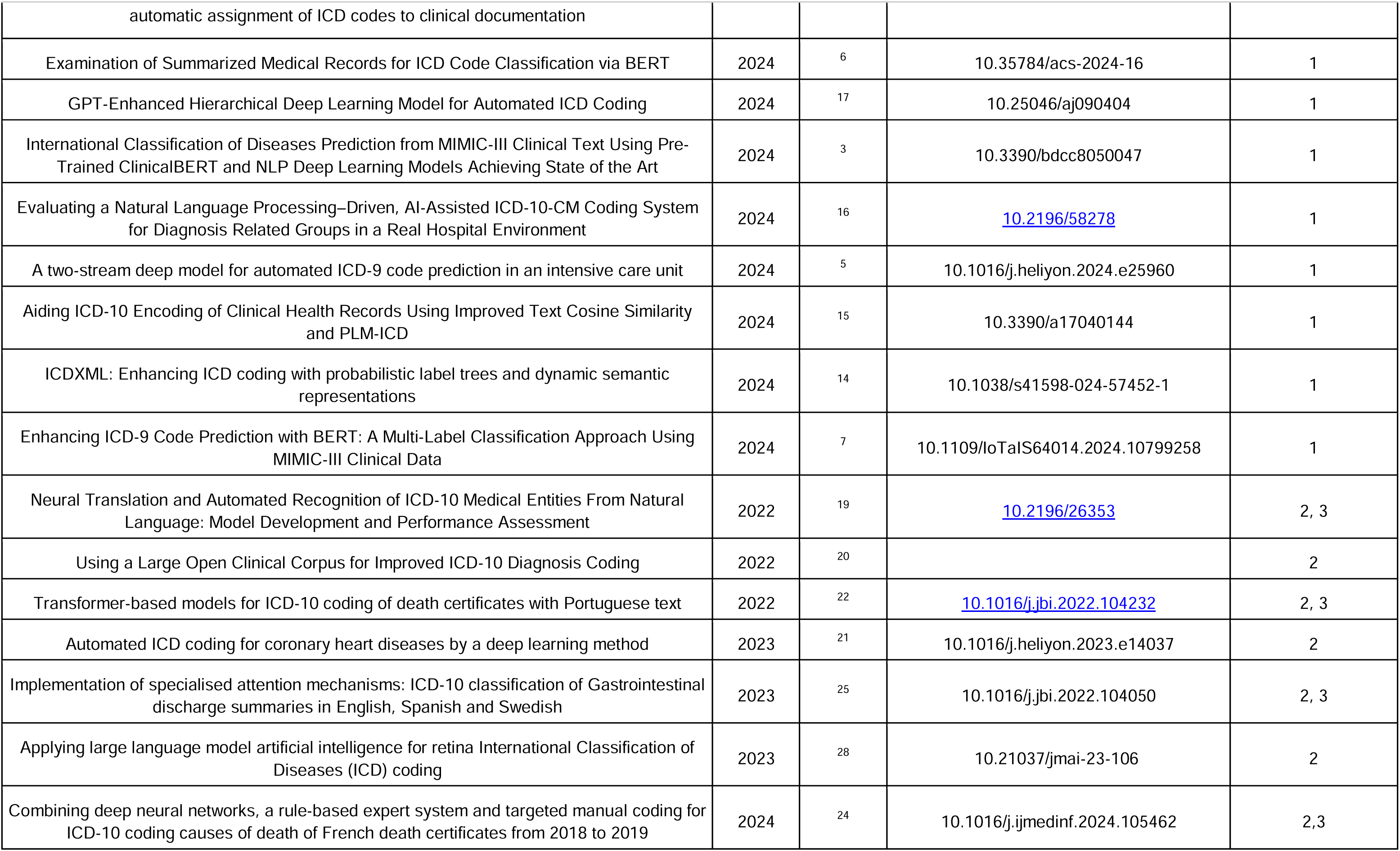
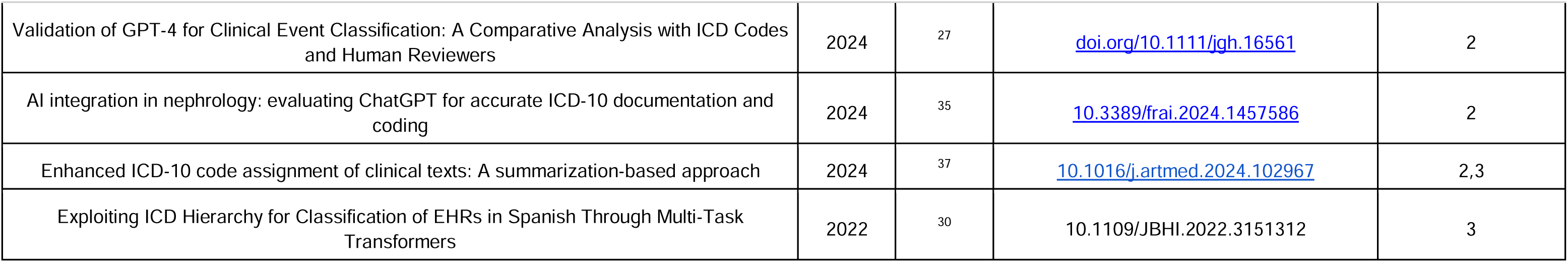
Overview of Selected Studies on Automated ICD Coding Using Large Language Models.

**Table 2:**
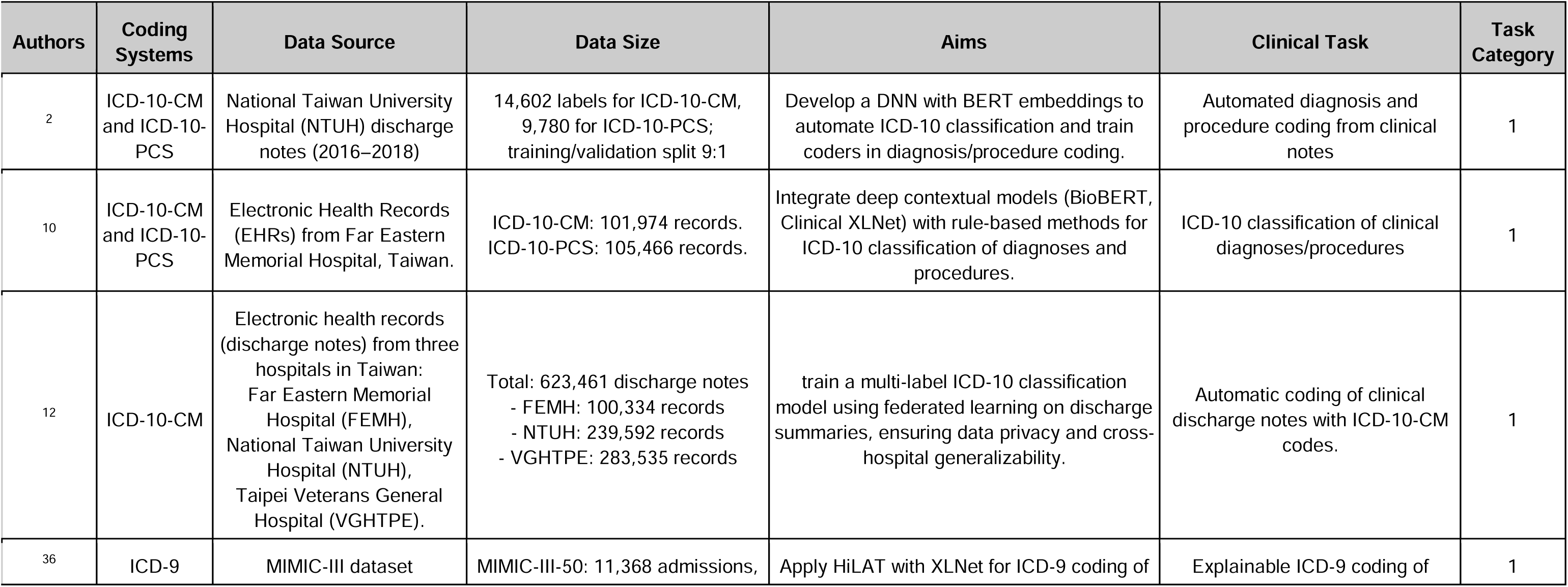
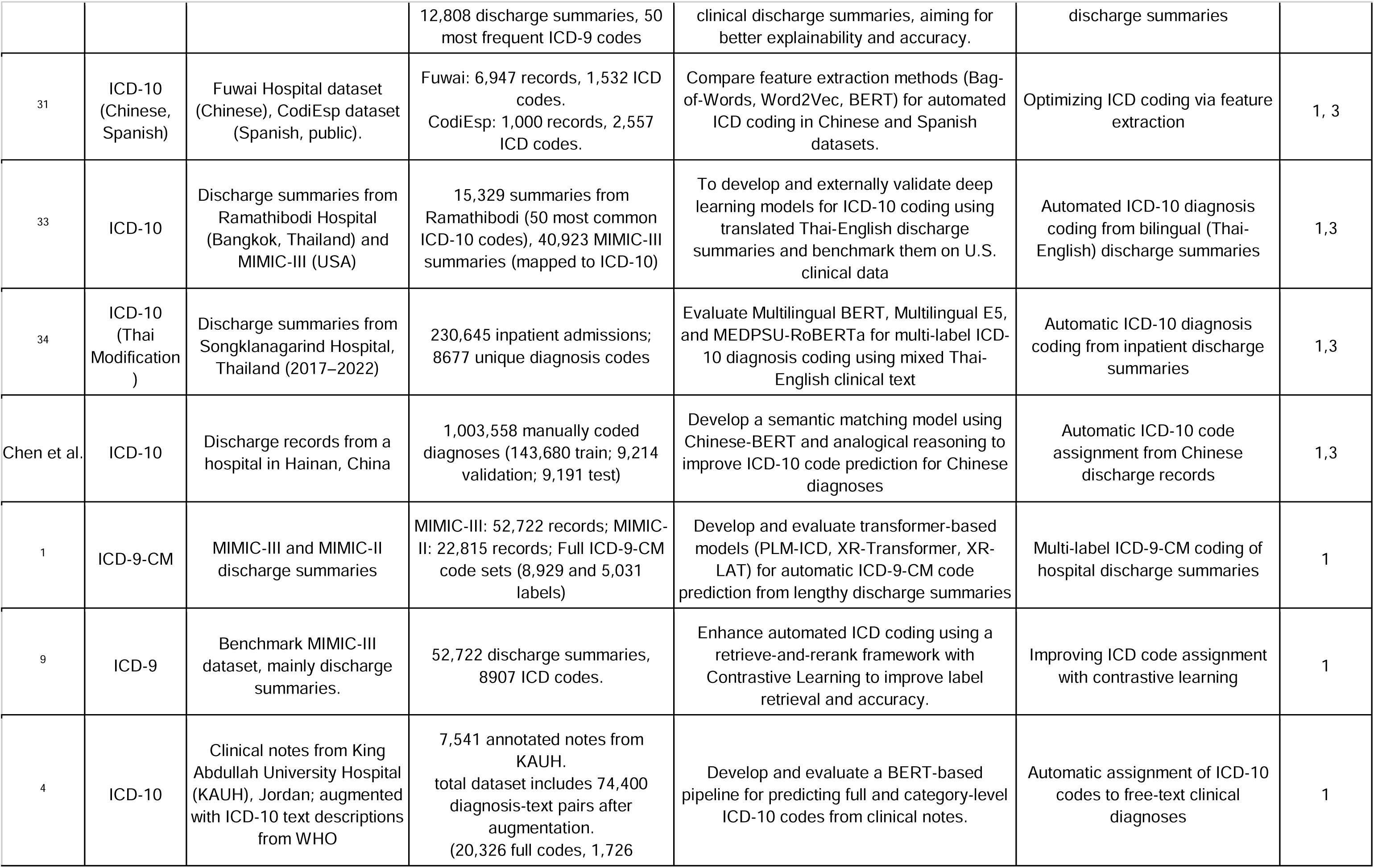
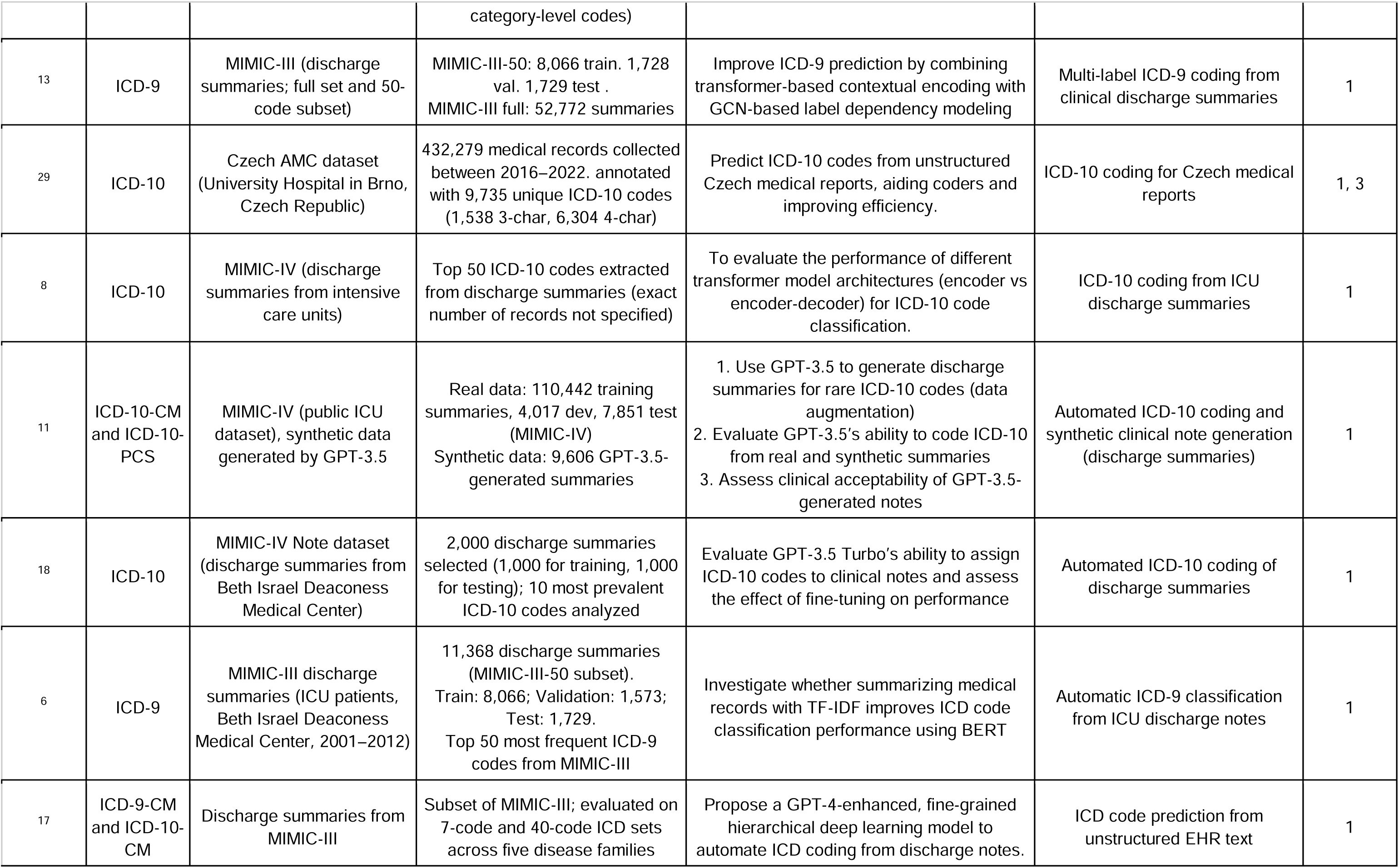
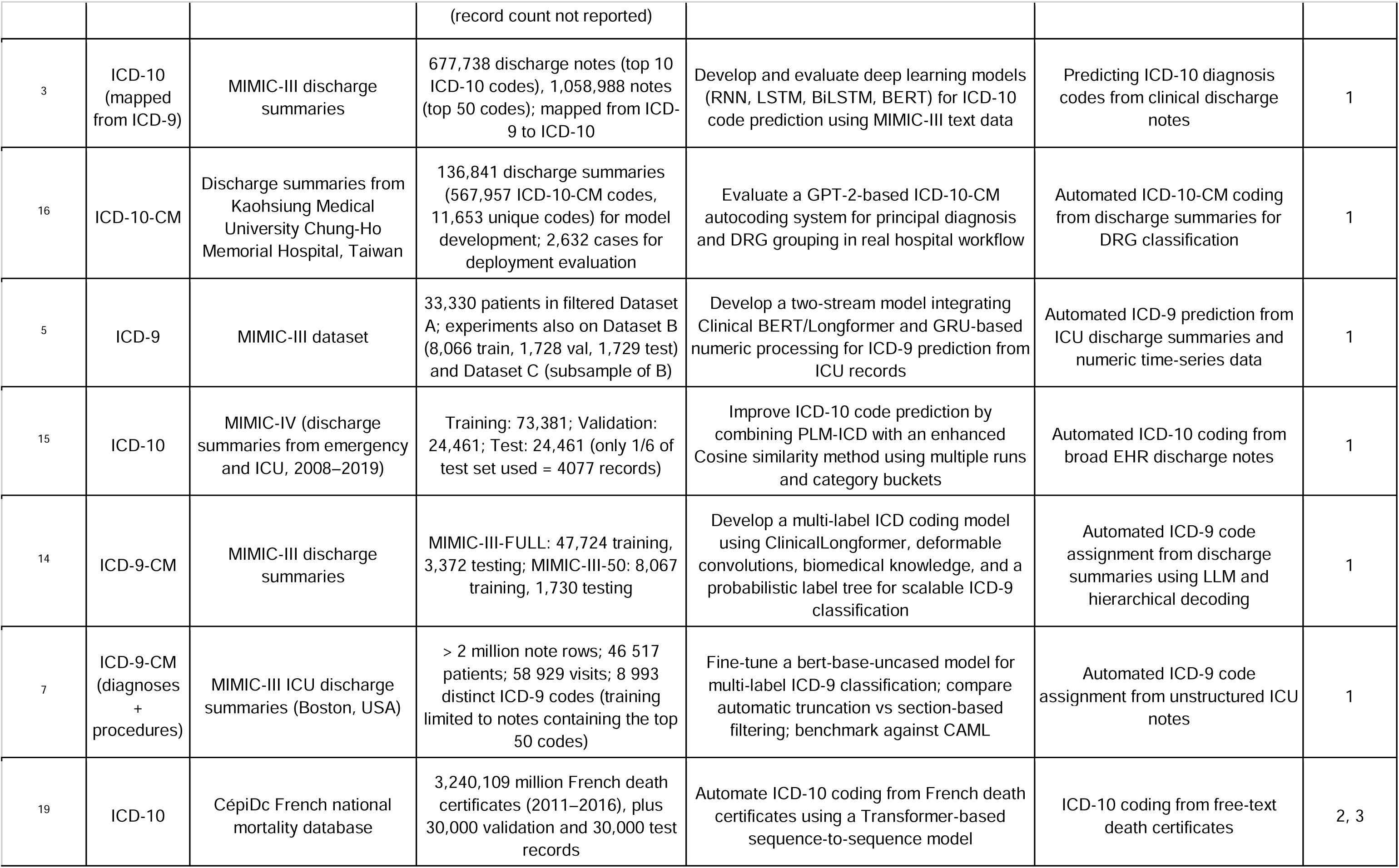
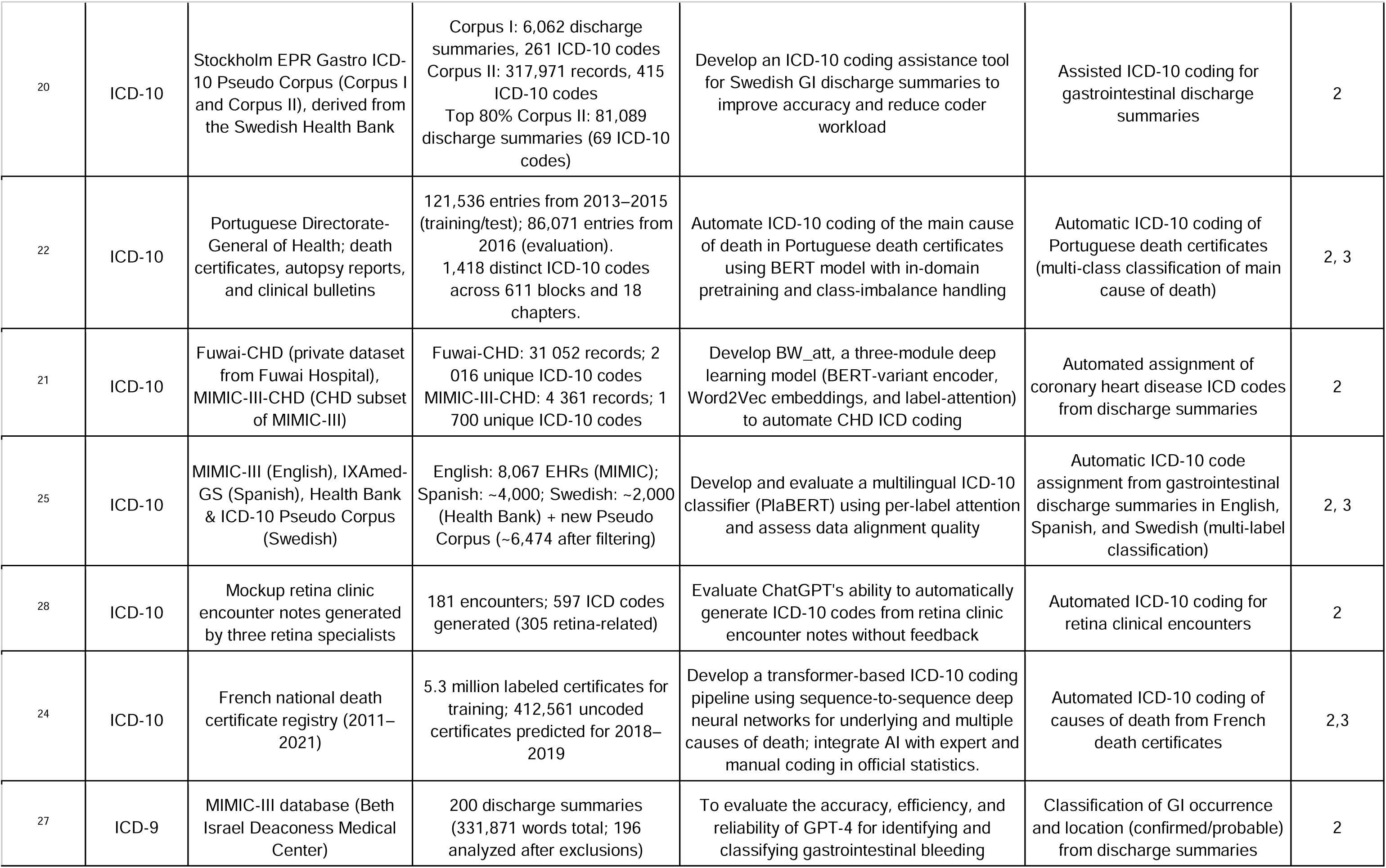
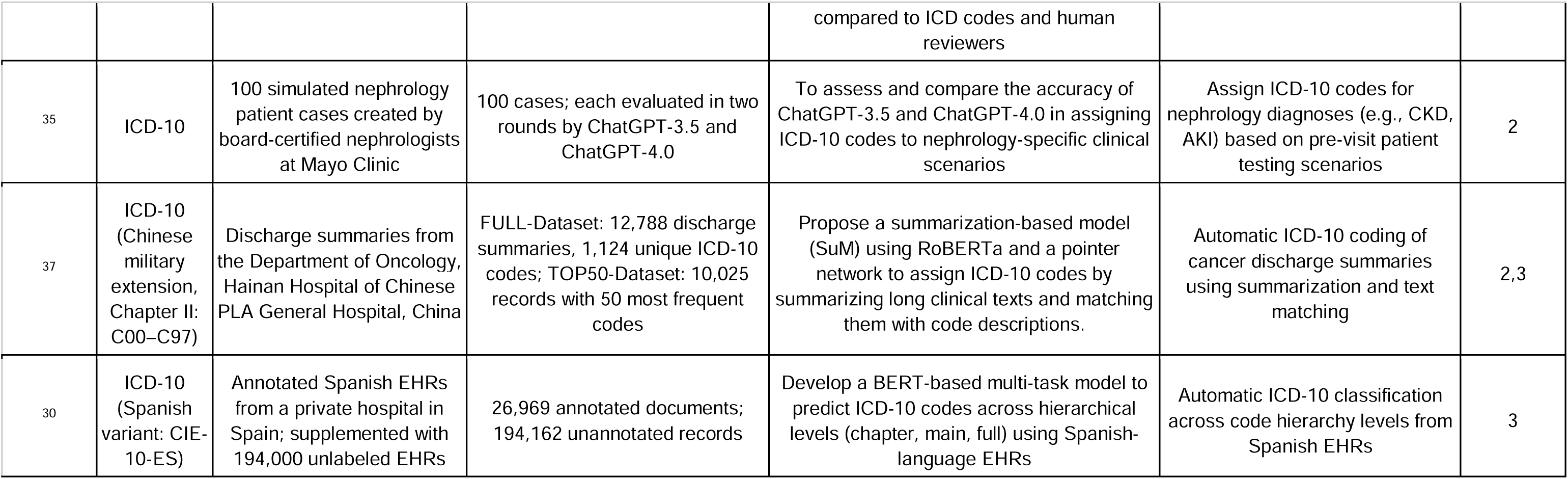
Overview of Clinical Tasks and Model Objectives in ICD-10 Coding Studies Using LLMs.

**Table 3:**
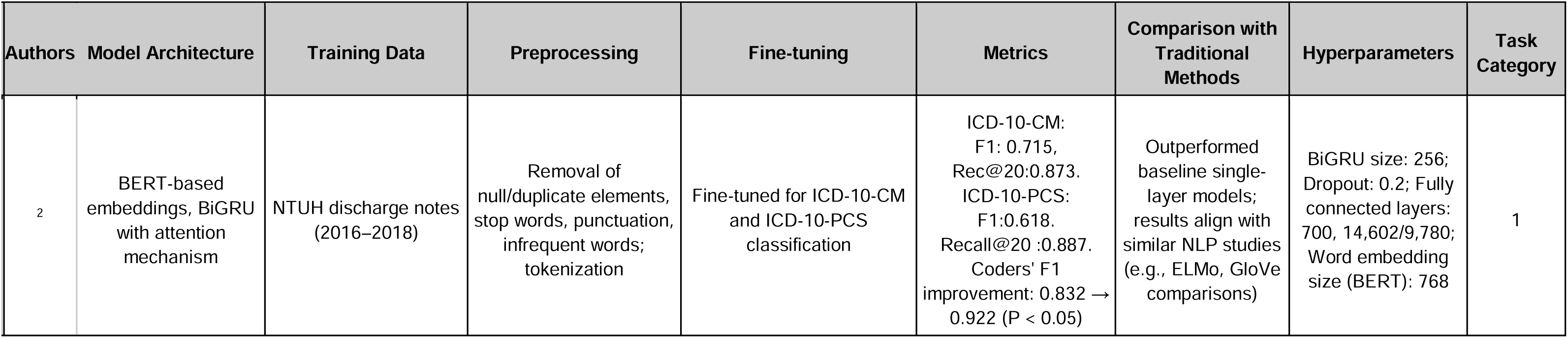
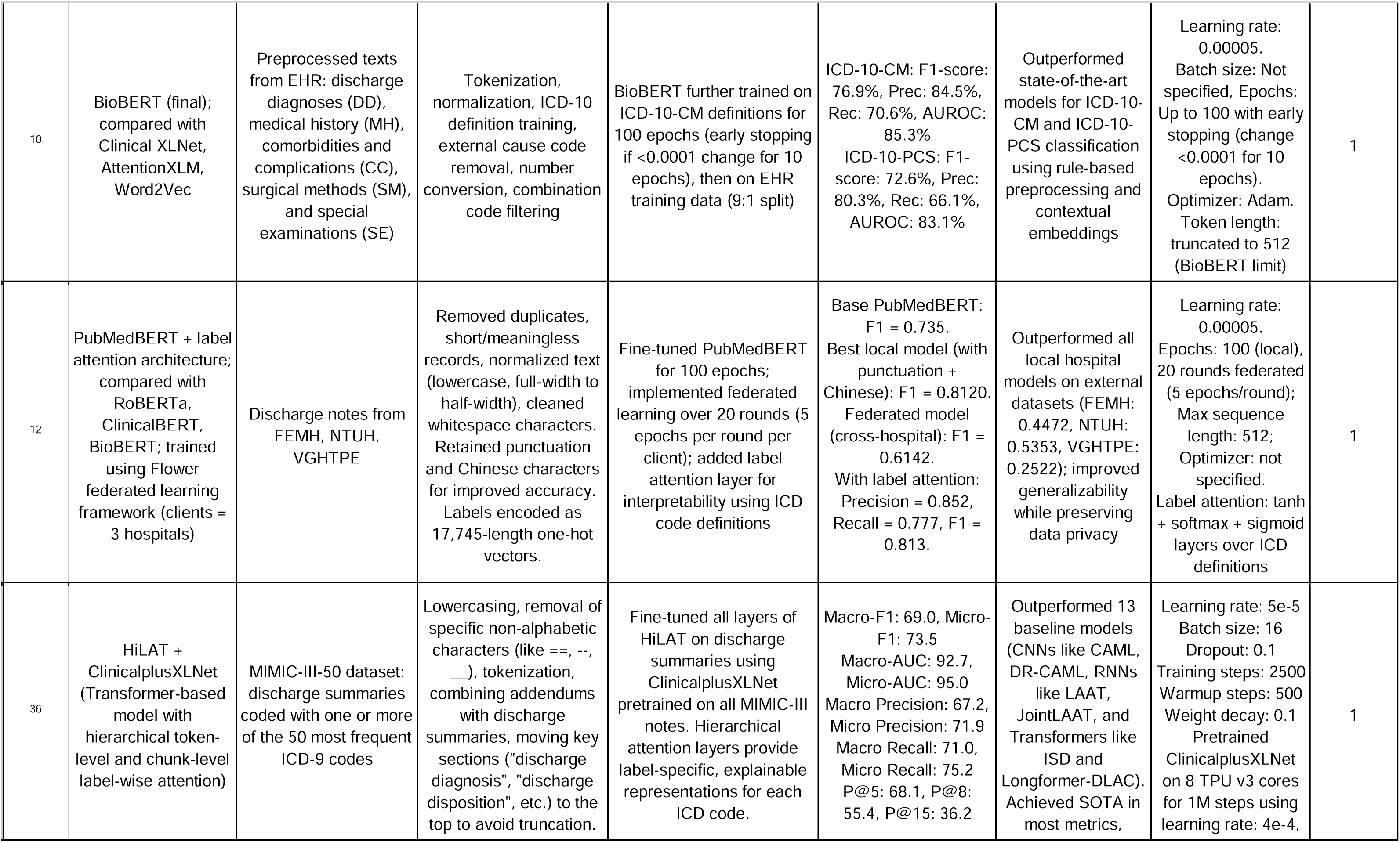
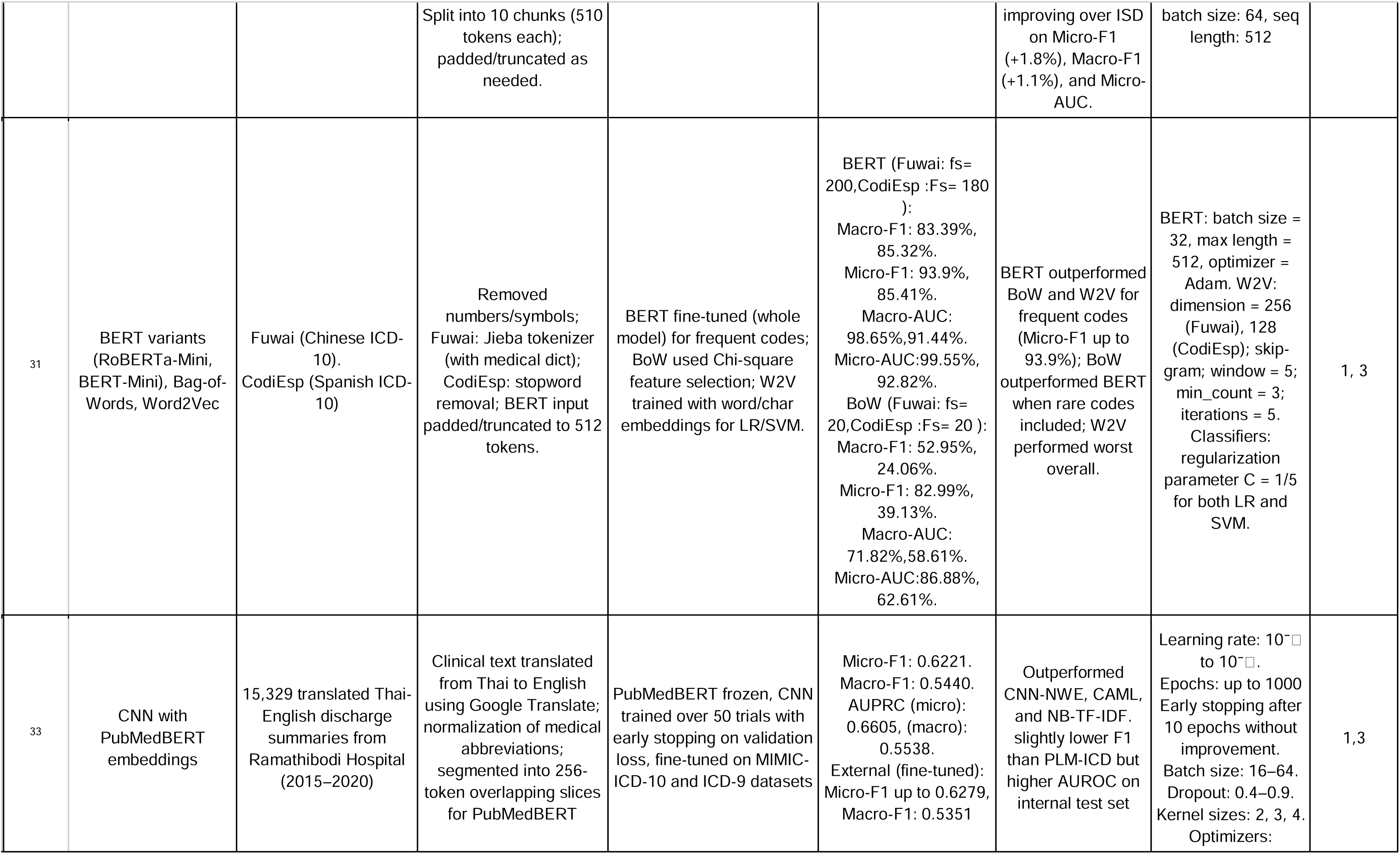
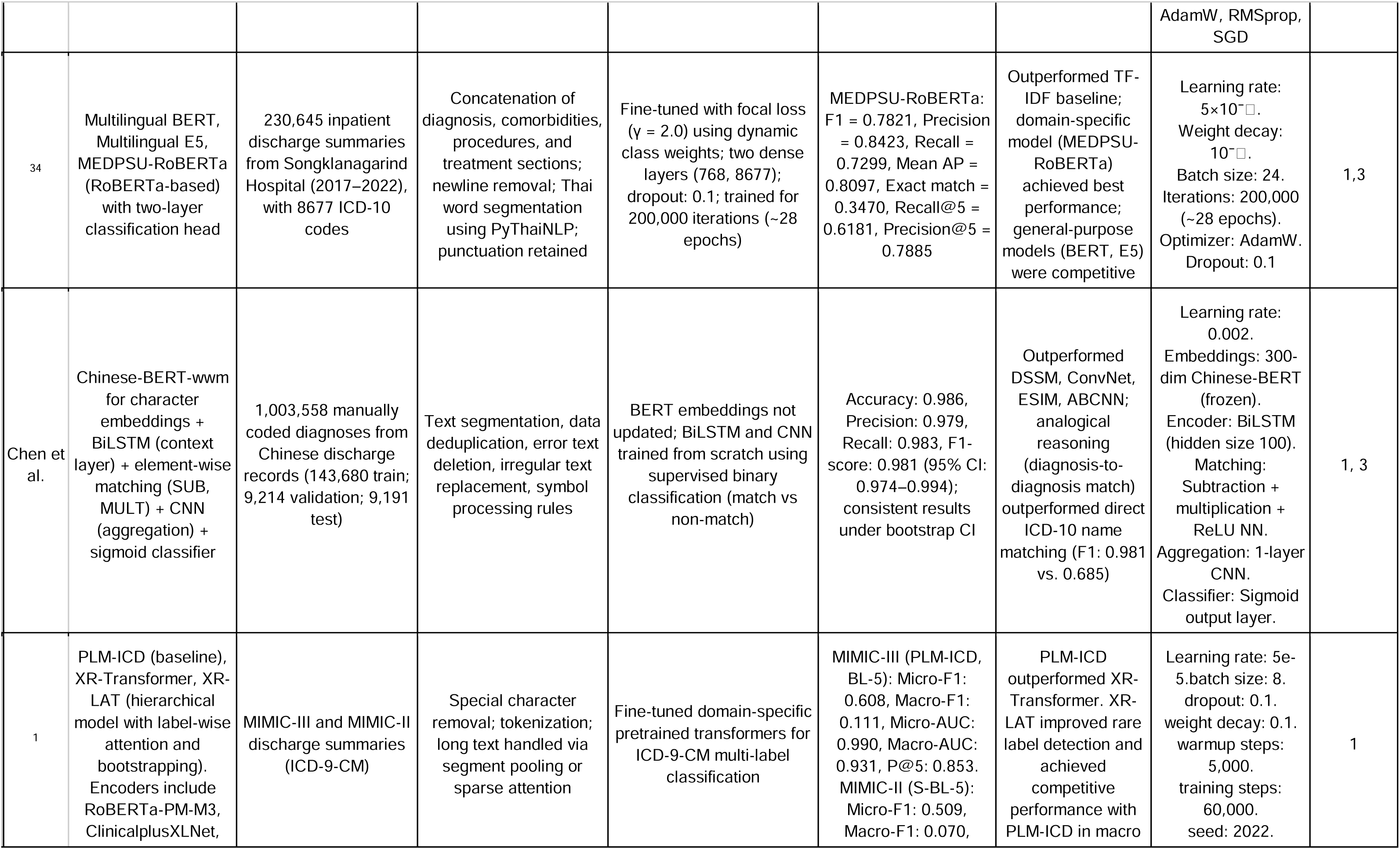
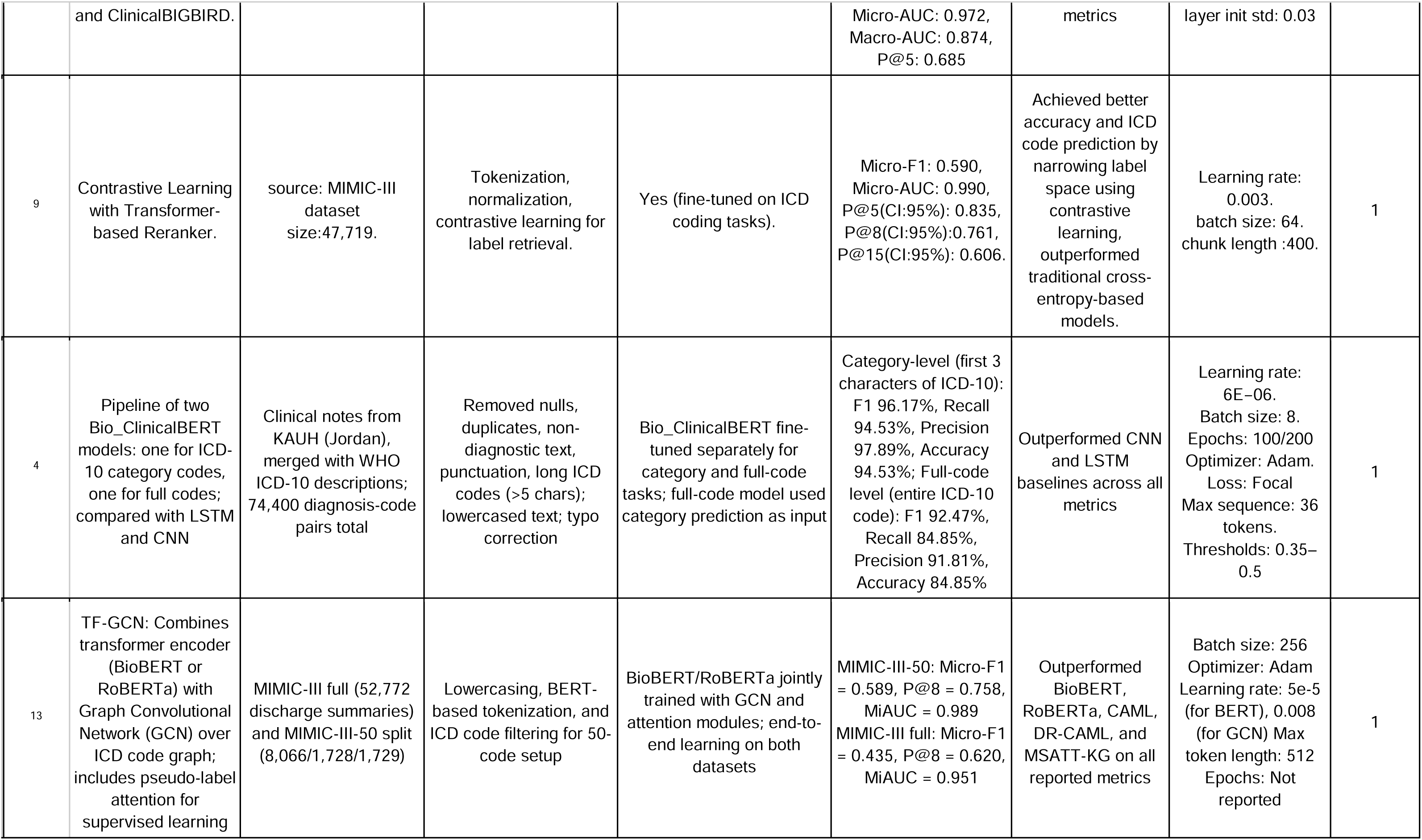
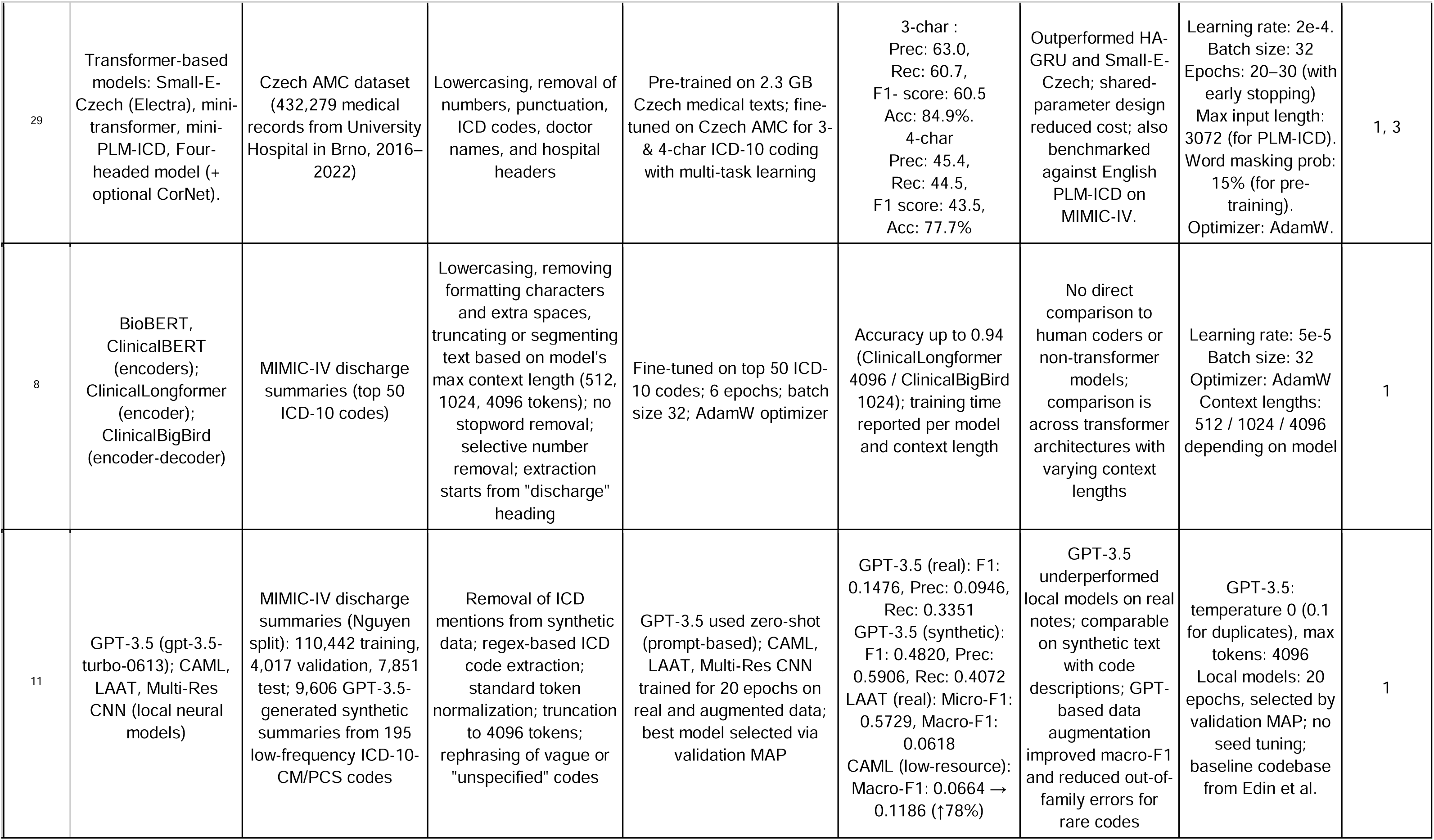
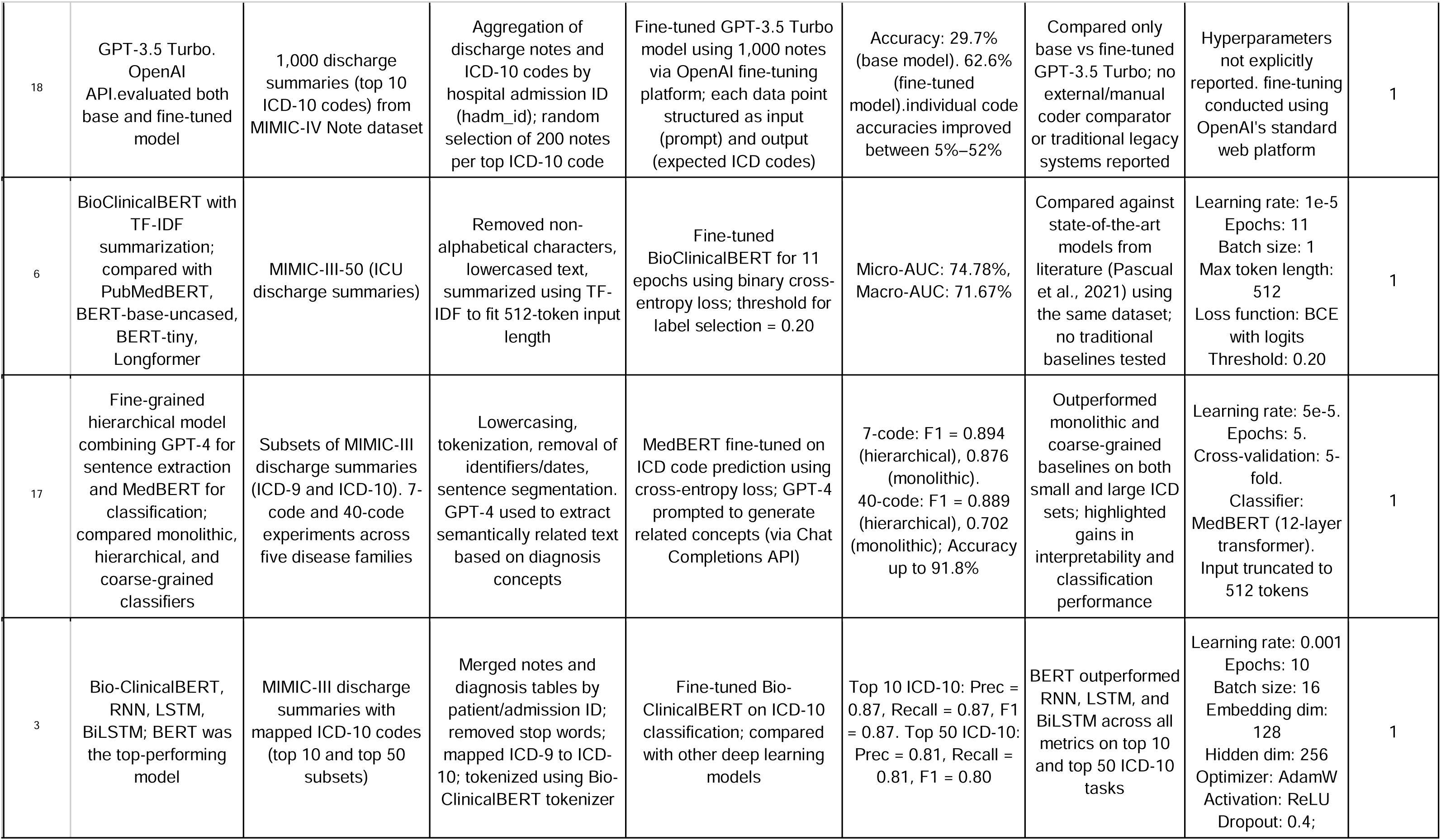
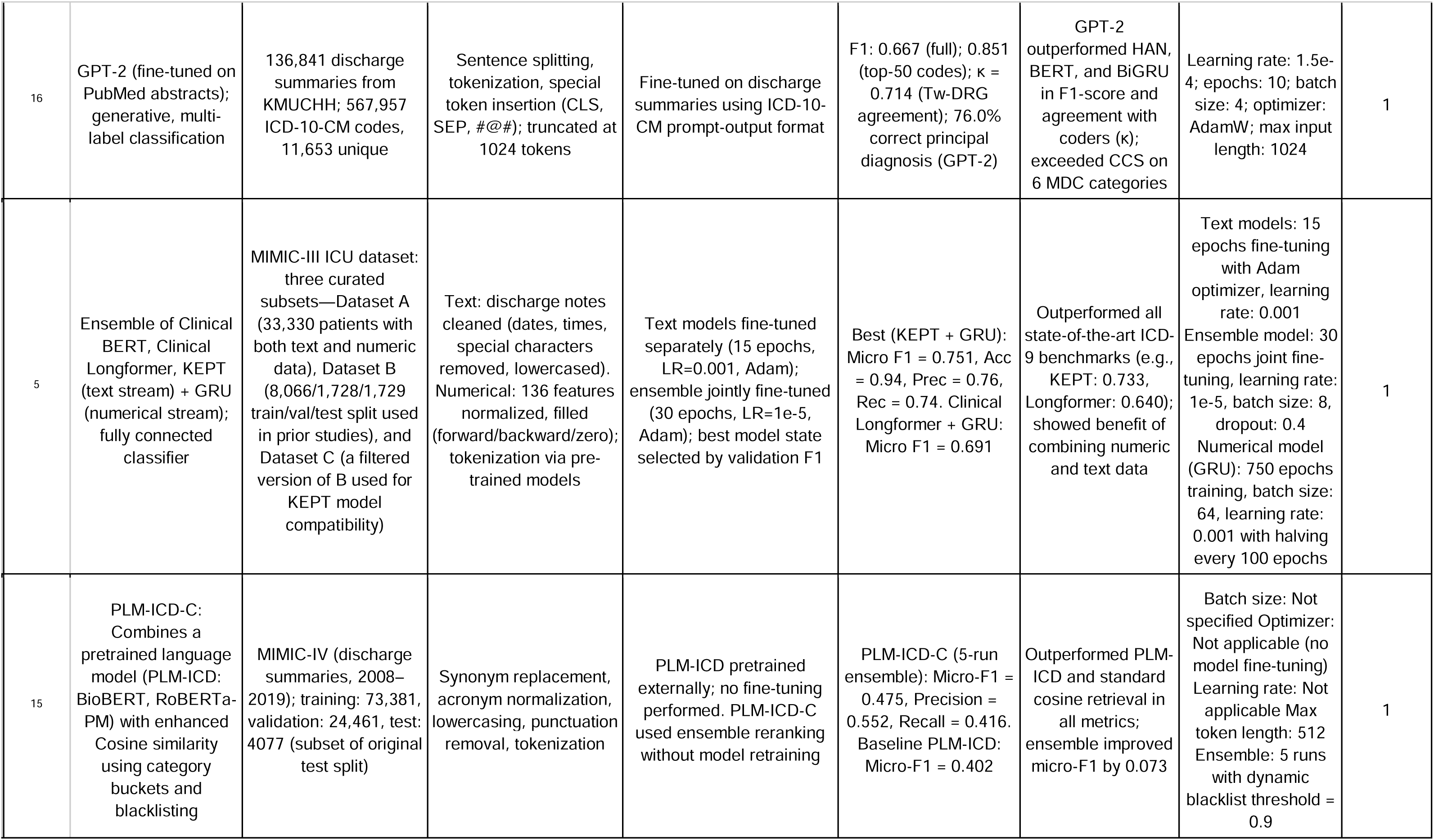
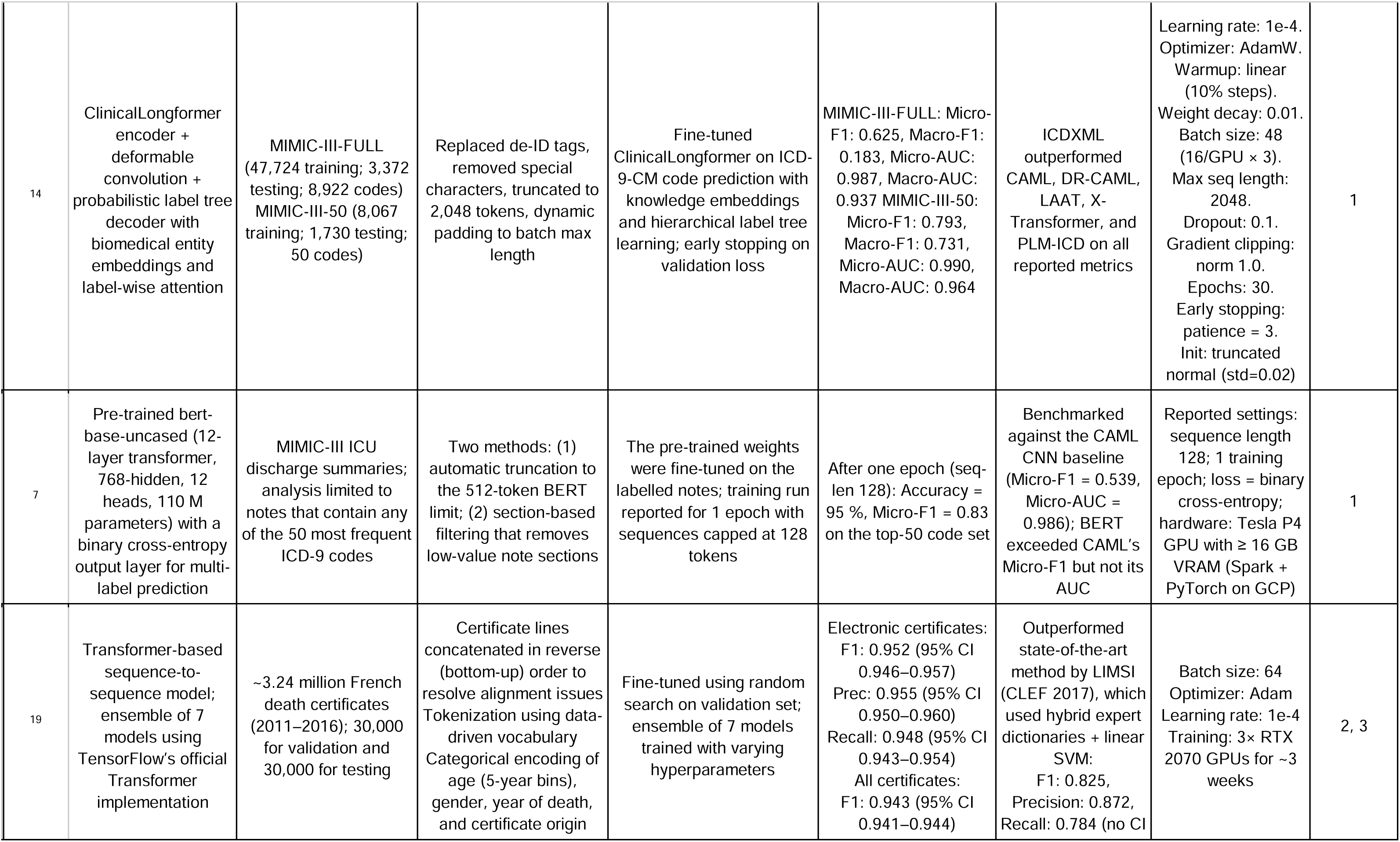
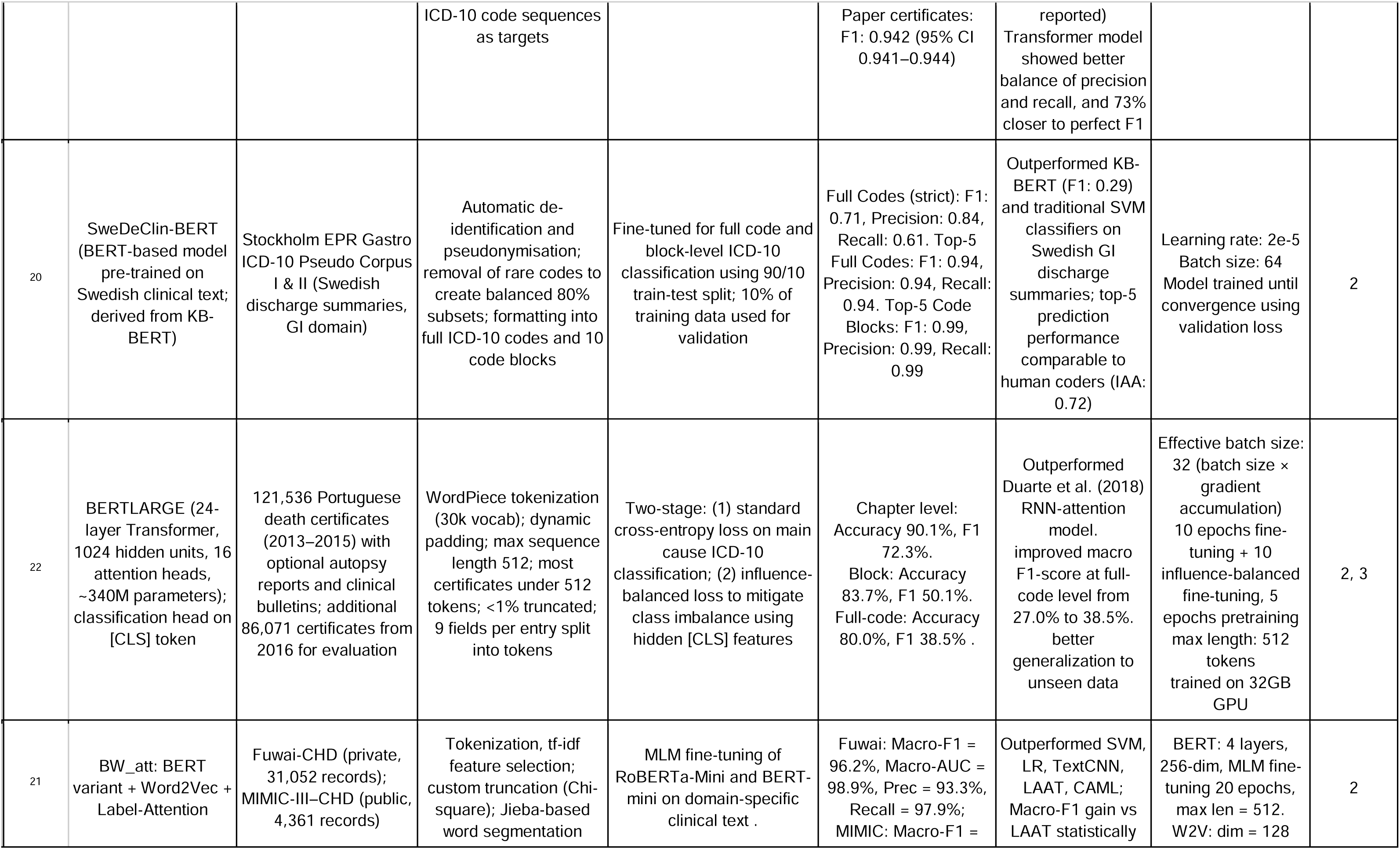
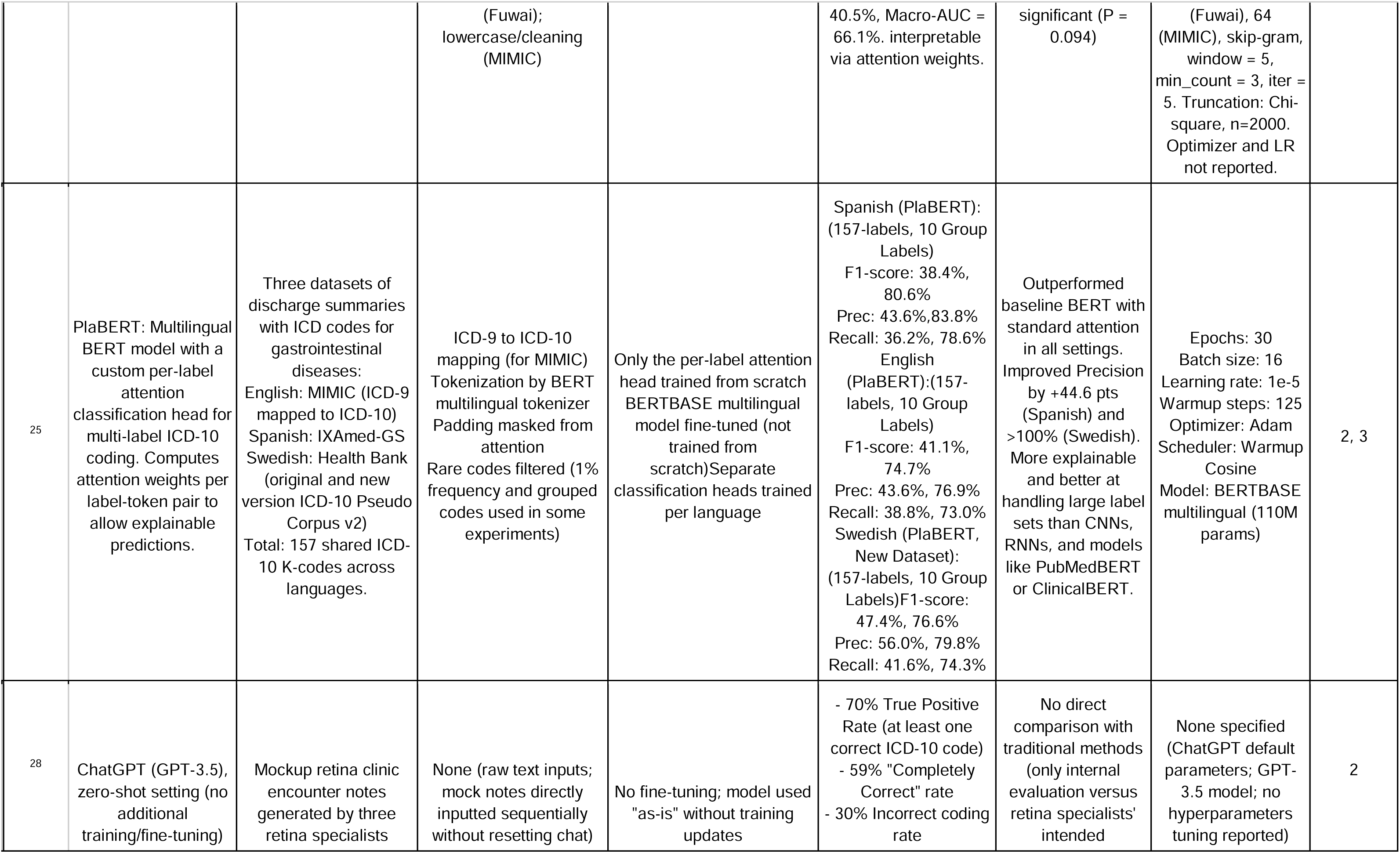
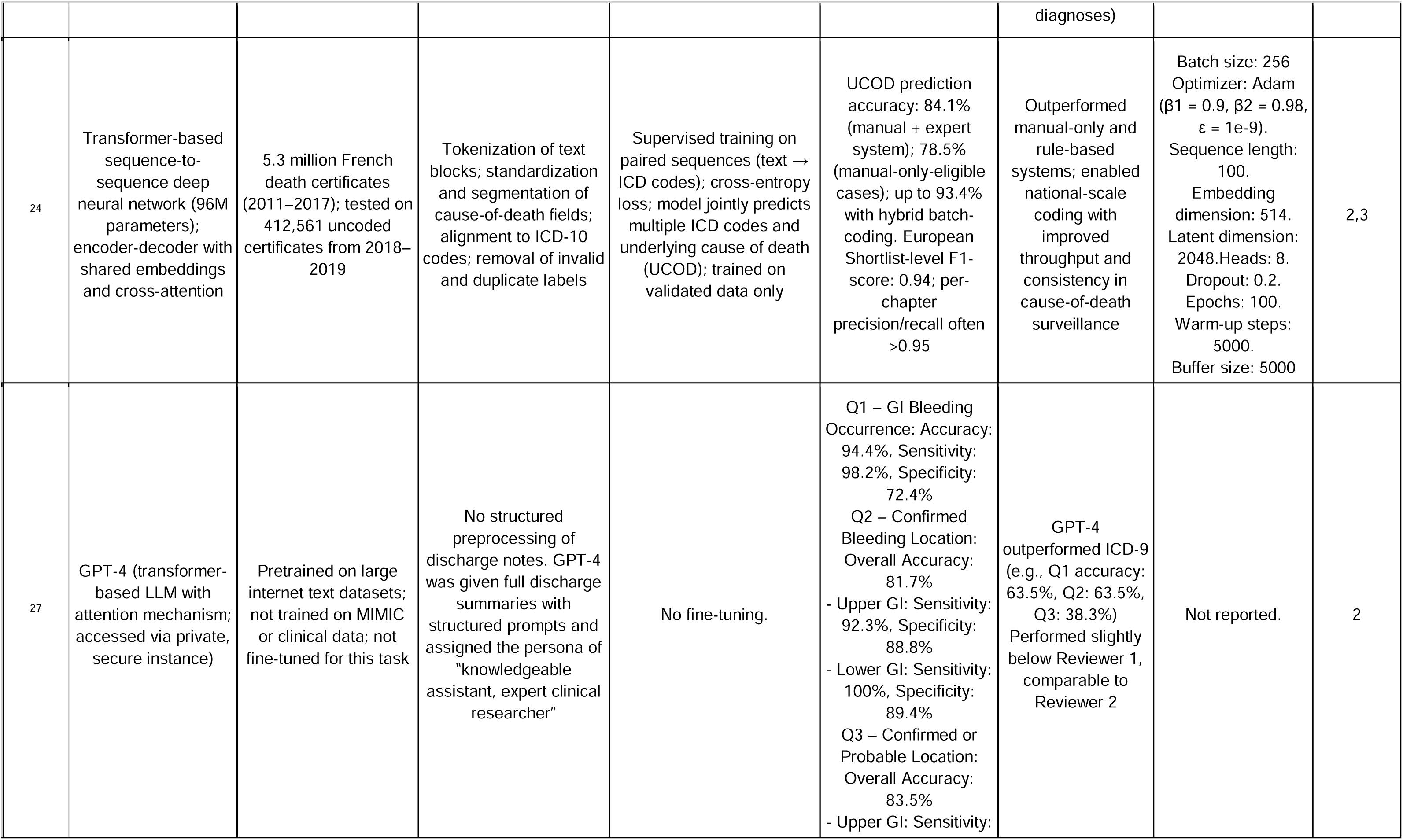
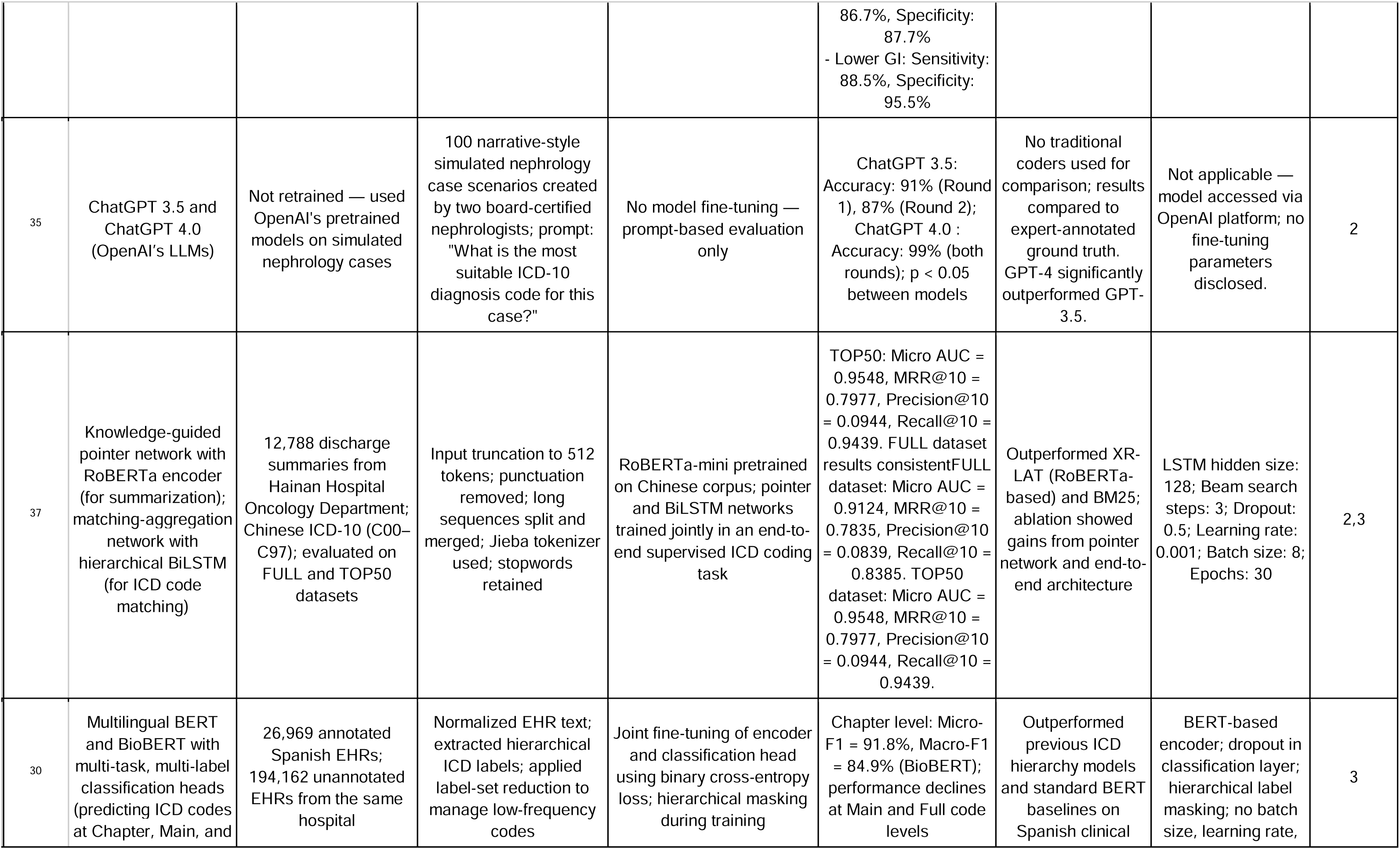
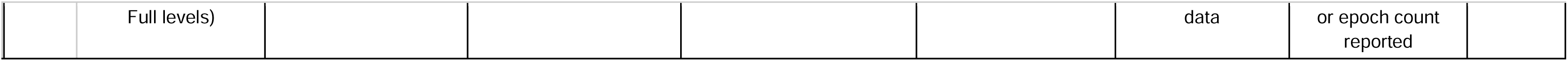
Technical Implementation and Performance Comparison of LLM Approaches for ICD-10 Coding.

**Table 4:** Strengths and Limitations of Reviewed Studies on LLM-Based ICD-10 Coding.

| Authors | Strengths | Limitations | Category |
| --- | --- | --- | --- |
| 2 | - Achieved F1-score of 0.715 for ICD-10 Clinical Modification (CM) codes and 0.618 for ICD-10 Procedure Coding System (PCS) codes.<br>- Integrated an auto-coding and training system, improving coder accuracy.<br>- Applied attention mechanisms to highlight relevant text for ICD coding.<br>- Evaluated on real-world hospital data (National Taiwan University Hospital, 2016–2018). | - Limited to a single medical center (Taiwan), making generalizability uncertain.<br>- Struggles with combination codes, leading to potential classification errors.<br>- Did not reduce coding time, as coders still took ~30 minutes per case.<br>- No validation on multilingual datasets or international ICD coding guidelines. | 1 |
| 10 | -BioBERT outperformed other models (F1: 0.701).<br>-selected for final model. Rule-based preprocessing (definition training, code removal, number conversion, combination filtering) improved F1 (ICD-10-CM: 0.749→0.769; PCS: 0.670→0.726).<br>-Used multiple EHR sections (medical history, comorbidities, surgical notes, exams) for richer context.<br>-High AUROC (CM: 0.853, PCS: 0.831), enhancing reliability.<br>-Leverages expert coding rules for improved interpretability and performance. | -Single-center data from Taiwan; limited generalizability.<br>-No external validation; needs replication across settings.<br>-Underperformance in rare ICD codes and complex multilabel cases.<br>-Computationally intensive due to large model size and rule-based layers.<br>-PCS prediction still less accurate than CM due to sparse/fragmented input data. | 1 |
| 12 | – Demonstrated federated learning can protect data privacy while improving model generalizability across hospitals.<br>– PubMedBERT outperformed ClinicalBERT, BioBERT, and RoBERTa (F1: 0.735).<br>– Best local model (with punctuation and Chinese): F1: 0.8120.<br>– Federated model achieved weighted F1: 0.6142 across hospitals.<br>– Label attention architecture enhanced interpretability without major performance loss (F1: 0.813).<br>– Showed punctuation and Chinese characters improve model performance. | – Data limited to three hospitals in Taiwan; generalizability beyond this setting is untested.<br>– Federated training required ~1 week, longer than local training (~2 days).<br>– VGHTPE hospital model underperformed due to label distribution mismatch.<br>– Did not address primary vs. secondary ICD code ordering in predictions.<br>– Translation of Chinese characters (for preprocessing) may lack linguistic nuance. | 1 |
| 36 | - HiLAT achieves state-of-the-art performance in F1-score and AUC for ICD-9 code prediction on MIMIC-III-50.<br>- Introduces hierarchical label-wise attention, enabling label-specific explainability by highlighting influential text.<br>- Uses ClinicalplusXLNet, an XLNet-based model pretrained on all MIMIC-III notes, which outperforms CNN/RNN and BERT-based baselines.<br>- Better suited for long clinical documents compared to standard BERT due to XLNet's flexible input handling. | - Evaluation limited to ICD-9 codes; generalizability to ICD-10 or other coding systems remains untested.<br>- Trained and fine-tuned using 8 TPU v3 cores, making it computationally intensive and potentially impractical for many clinical settings.<br>- The model is only tested on MIMIC-III, limiting external validity to other healthcare systems.<br>- Focused exclusively on the 50 most frequent ICD codes, reducing effectiveness for rare or fine-grained diagnoses. | 1 |
| 31 | - Comprehensive feature extraction comparison across multiple machine learning models.<br>- Fine-tuned BERT models perform best for frequent codes (Micro-F1: 93.9% on Chinese data, 85.4% on Spanish data).<br>- Bag-of-Words (BoW) is optimal for infrequent codes, providing better interpretability.<br>- Analyzes multiple datasets (Chinese and Spanish clinical records), improving generalizability.<br>- Strong evaluation framework using multiple frequency thresholds to determine the best extraction method per task. | - Limited dataset size (Chinese dataset: 6947 records, Spanish dataset: 1000 records), affecting robustness.<br>- Only ICD-10 codes tested, lacking validation on ICD-9 or ICD-PCS.<br>- Findings may not generalize to datasets with different languages or healthcare systems.<br>- Feature extraction results are highly dependent on frequency threshold selection, making reproducibility complex. | 1, 3 |
| 33 | – Developed and validated CNN-PubMedBERT and PLM-ICD models for ICD-10 coding from Thai-English discharge summaries.<br>– External validation on MIMIC-ICD-10 and ICD-9 datasets demonstrated cross-setting applicability.<br>– Addressed input length limitations of BERT by segmenting long discharge texts. | – Limited to the 50 most frequent ICD-10 codes, reducing coverage of rare diagnoses.<br>– Source data (Ramathibodi Hospital) had mixed Thai-English notes, requiring translation which may introduce errors.<br>– Discharge summaries may omit comorbidities, reducing label completeness. | 1,3 |
|  | – Integrated translation, abbreviation normalization, and segment tokenization for robust text processing.<br>– Released model code publicly for reproducibility. | – Fine-tuning improved performance, but generalizability to other languages and healthcare systems remains untested.<br>– Did not incorporate structured data or temporal relationships from patient histories. |  |
| 34 | – Large dataset (230,645 records, 8677 codes) from real-world inpatient care.<br>– Compared general and domain-specific models; MEDPSU-RoBERTa achieved top F1 and MAP.<br>– Used focal loss and dynamic weighting to address code imbalance.<br>– Handled mixed Thai-English text with tailored preprocessing. | – Focused on 50 most frequent codes; limited rare code coverage.<br>– Translation and segmentation may introduce noise.<br>– Lacked structured or historical data. – No external validation beyond one hospital. | 1,3 |
| 32 | – Achieved high performance (F1: 0.981) using analogical reasoning with BERT embeddings.<br>– Outperformed DSSM, ConvNet, ESIM, and ABCNN on ICD-10 coding.<br>– Validated on over 1 million real-world Chinese discharge diagnoses.<br>– Demonstrated robustness via both 95% CI and bootstrap CI. | – Limited to Chinese text; not evaluated on multilingual or cross-country datasets.<br>– Does not incorporate full clinical context beyond diagnoses.<br>– Computationally slower than simpler models.<br>– Data not publicly shareable; limits external reproducibility. | 1, 3 |
| 1 | – Evaluated multiple transformer-based architectures on full-label ICD-9-CM datasets.<br>– PLM-ICD and XR-LAT models showed strong performance on both frequent and rare codes.<br>– Demonstrated robust performance across two real-world datasets (MIMIC-III and MIMIC-II).<br>– XR-LAT improved macro-F1 and macro-AUC using recursive bootstrapping and label-aware training. | – Focused only on ICD-9-CM; not tested on ICD-10 or multilingual settings.<br>– No real-time or workflow integration studies included.<br>– Evaluation limited to two US-based datasets; generalizability to other systems untested. | 1 |
| 9 | - Contrastive Learning (CL) improves label retrieval, reducing irrelevant ICD codes before ranking.<br>- Achieves 0.590 Micro-F1 and 0.990 Micro-AUC, outperforming traditional models.<br>- Addresses ICD code imbalance by refining the label space.<br>- Uses a Transformer variant with lower complexity, reducing memory consumption for long clinical texts.<br>- Publicly available source code, facilitating reproducibility. | - Tested only on MIMIC-III (ICD-9); not validated on ICD-10 or real-world hospital data.<br>- Hallucinations in predictions may still occur despite improved label retrieval.<br>- Contrastive Learning requires careful tuning of hyperparameters for optimal performance.<br>- Complex two-stage approach requires additional computational resources compared to simpler deep learning models. | 1 |
| 4 | – Developed and compared LSTM baseline and BERT-based pipeline for ICD-10 code prediction.<br>– Achieved strong performance with F1-score of 92.47% and precision of 91.81% (Pipeline BERT). | – Data limited to a single institution in Jordan; generalizability untested.<br>– Focused on single-label classification; does not support multi-label ICD coding.<br>– Small dataset size compared to label diversity (20,326 full codes). | 1 |
|  | – Used domain-specific pretraining (Bio_ClinicalBERT on MIMIC-III) to boost performance.<br>– Addressed class imbalance using focal loss. | – No evaluation on multilingual datasets or real-time deployment. |  |
| 13 | – TF-GCN achieved state-of-the-art results on both MIMIC-III-50 and full datasets.<br>– Effectively integrated textual semantics (via BioBERT/RoBERTa) and code relationships (via GCN).<br>– Introduced pseudo-label attention to enhance supervision during training | – Model performance decreased on the full dataset compared to the 50-code subset.<br>– Study limited to English-language ICD-9 and MIMIC-III data.<br>– Did not report macro-F1 or conduct external validation | 1 |
| 29 | - Designed specifically for Czech medical reports, ensuring strong adaptability to the local healthcare system.<br>- Utilizes compact transformer architectures (e.g., Four-headed model), significantly reducing computational and memory requirements — suitable for deployment in resource-limited hospitals.<br>- Introduces a novel "Four-headed" multi-task model that improves learning efficiency and parameter sharing.<br>- Achieves performance comparable to state-of-the-art English models (like PLM-ICD) on the MIMIC-IV benchmark, despite being smaller and trained on less data. | - Limited to Czech-language reports; generalization to other languages or healthcare systems not tested.<br>- Predicts ICD-10 codes up to the 4-character level (not full codes), requiring manual review or completion by human coders.<br>- The dataset (Czech AMC) is proprietary and not publicly available, restricting external validation or replication. | 1, 3 |
| 8 | - Benchmarks multiple transformer architectures (BioBERT, ClinicalBERT, ClinicalLongformer, ClinicalBigBird) using real ICU discharge data (MIMIC-IV).<br>- Evaluates the effect of context length on model performance.<br>- Achieves high accuracy (up to 0.94) with long-context models.<br>- Includes training time analysis to inform model choice in resource-constrained settings. | - Limited to the top 50 ICD-10 codes; does not generalize to the full code set.<br>- No comparison with human coders or traditional machine learning methods.<br>- Accuracy gains from increased context length are modest and come at significantly higher computational cost.<br>- No external validation on datasets outside MIMIC-IV. | 1 |
| 11 | - GPT-3.5 can generate discharge summaries that improve performance for rare ICD codes when used for data augmentation.<br>- Introduced a novel evaluation of GPT for synthetic data generation and direct coding.<br>- Real clinical evaluation by physicians adds credibility to acceptability assessments.<br>- Open access data and code provided for reproducibility. | - GPT-3.5 underperforms on real clinical notes without prompt-based guidance.<br>- Synthetic notes often lack coherence, informativeness, and narrative realism.<br>- Clinical note generation includes hallucinated or irrelevant details.<br>- Evaluators reported poor acceptability for clinical deployment. | 1 |
| 18 | - Demonstrated fine-tuning improved ICD-10 assignment accuracy from 29.7% to 62.6%. | - Limited to 10 most common ICD-10 codes; generalizability to rarer codes untested. | 1 |
|  | - Used real-world de-identified EHR notes (MIMIC-IV Note).<br>- Highlighted fine-tuning as a resource-efficient alternative to developing custom models.<br>- Showed cloud-based AI can enhance healthcare administrative tasks. | - No direct comparison with human coders or legacy auto-coding systems.<br>- Fine-tuning sample size arbitrary (100 notes per code); may not maximize model performance.<br>- Hyperparameters and detailed fine-tuning settings not reported.<br>- External validation on other datasets missing. |  |
| 6 | - Proposed a novel approach combining TF-IDF summarization with BioClinicalBERT for ICD classification on MIMIC-III-50.<br>- Enables real-time ICD code classification with improved interpretability.<br>- Evaluated multiple BERT variants and summarization strategies.<br>- Focused on shorter, more readable summaries for clinical usability. | - Lower performance than state-of-the-art models (Macro-AUC 71.67%, Micro-AUC 74.78%).<br>- Summarization and classification were conducted independently, which may have introduced noise.<br>- Performance loss likely due to irrelevant info in summaries.<br>- Limited to top 50 ICD-9 codes only (not full code set). | 1 |
| 17 | - Achieved F1-score of 0.894 on a 7-code set and 0.889 on a 40-code set using a hierarchical classifier.<br>- Fine-grained coding strategy reduced classification complexity by predicting one ICD code per diagnosis.<br>- GPT-4 used to extract semantically related sentences from discharge notes, enriching classifier input.<br>- Hierarchical model design aligned with ICD code taxonomy to enhance interpretability and scalability. | - Dataset size and specific ICD code distributions were not reported.<br>- Label imbalance noted as a challenge; no direct mitigation methods implemented.<br>- No comparison with commercial systems or external validation performed. | 1 |
| 3 | - Achieved F1 score of 0.87 for top 10 ICD-10 code prediction and 0.80 for top 50 codes using Bio-ClinicalBERT.<br>- Outperformed previous studies using deep learning for ICD prediction on the same task.<br>- Compared multiple deep learning models (RNN, LSTM, BiLSTM, BERT) under the same experimental setup.<br>- Employed robust preprocessing and hyperparameter tuning to improve model performance. | - Faced computational limitations for training complex models on large datasets.<br>- MIMIC-III data complexity introduced issues like imbalanced classes and missing values.<br>- Models required extensive preprocessing and fine-tuning to mitigate overfitting and gradient issues.<br>- BERT and other transformer models required significant compute resources for training and tuning. | 1 |
| 16 | - Demonstrated GPT-2 outperforms HAN, BERT, and BiGRU in F1 and DRG agreement.<br>- Achieved $\kappa = 0.714$ with certified coders across six MDC groups.<br>- Evaluated in a real hospital setting using actual discharge summaries.<br>- Principal diagnosis prediction accuracy of 76.0%.<br>- Performance sustained across top-50 and full ICD-10-CM code sets. | - GPT-2 model incorrectly predicted 24.0% of principal diagnoses.<br>- Only one institution was used; external generalizability not tested.<br>- Does not handle procedure codes (ICD-10-PCS).<br>- No multilingual validation or non-Taiwan healthcare evaluation.<br>- Model and evaluation limited to discharge summaries only. | 1 |
| 5 | – Highest micro F1 (0.751) among ICD-9 models by combining text and numeric data.<br>– Improved detection of hard-to-predict codes (e.g., thrombocytopenia).<br>– Outperformed prior benchmarks (KEPT, Longformer) on standard datasets. | – Limited to ICU data from MIMIC-III; generalizability unclear.<br>– No external or multilingual validation.<br>– High training cost due to ensemble architecture. | 1 |
| 15 | – Improved micro-F1 score from 0.402 (PLM baseline) to 0.475 with the PLM-ICD-C ensemble.<br>– Combined transformer-based model with Cosine similarity search enhanced by category buckets and dynamic filtering.<br>– Required no additional model training or parameter tuning. | – Evaluation limited to one dataset (MIMIC-IV) and English-language ICD codes.<br>– Only 1/6 of the original test set used for evaluation (4,077 records).<br>– No external or multilingual validation. | 1 |
| 14 | – Integrates LLM, biomedical knowledge, and probabilistic label trees for improved ICD coding.<br>– Outperforms prior models (CAML, DR-CAML, LAAT, PLM-ICD) on both frequent and rare codes.<br>– Achieves state-of-the-art results on MIMIC-III-50 and MIMIC-III-FULL datasets. | – Focused only on English-language ICD-9-CM data.<br>– Evaluation limited to MIMIC-III; lacks external validation.<br>– Model complexity and training time not evaluated against deployment constraints. | 1 |
| 7 | – Achieved 95 % evaluation accuracy and Micro-F1 0.83 after a single fine-tuning epoch on the 50 most frequent ICD-9 codes – Tackles BERT's 512-token limit by applying automatic truncation and section-based filtering during preprocessing – Draws on MIMIC-III's > 2 million patient-note rows as training text | – Model still below the CAML benchmark (Micro-AUC 0.91) despite higher F1 – Evaluation limited to the top-50 ICD-9 codes; rare codes are deferred to future work – Authors note that learning-rate, batch size and longer sequence lengths (256–512 tokens) need further tuning and testing | 1 |
| 19 | – High performance (F1 = 0.952), surpassing prior methods (F1 = 0.825)<br>– Trained on ~3.24M real-world death certificates<br>– End-to-end Transformer model, no manual features<br>– Well-calibrated scores enable semi-automated coding<br>– Expert validation and Turing test support reliability | – Lower F1 on paper forms due to missing data<br>– Poorer performance on rare ICD-10 categories<br>– Evaluation labels may include human errors<br>– Limited interpretability<br>– Not tested in other languages<br>– Needs large labeled datasets | 1 |
| 20 | - High accuracy for ICD-10 coding, particularly in the gastrointestinal domain.<br>- The top-5 prediction approach helps medical coders by providing a shortlist of likely ICD-10 codes.<br>- Uses a domain-specific BERT model (SweDeClin-BERT), improving performance over general NLP models.<br>- Based on large, real-world clinical data from the Swedish Health Bank.<br>- De-identified and pseudonymized data, allowing for research access. | - Limited to Swedish clinical data, reducing applicability to other languages and healthcare systems.<br>- ICD code imbalance: Some rare ICD-10 codes have very few examples, impacting prediction accuracy.<br>- Corpus I is too small for reliable training, leading to poor performance. | 2 |
| 22 | - Outperformed prior GRU-based model on all ICD levels.<br>- Effective in low-resource, non-English (Portuguese) setting.<br>- Novel in-domain pretraining and class imbalance handling.<br>- Generalized well to unseen 2016 data. | - Performance drops at more detailed ICD levels (Full-code F1 = 38.5%).<br>- Lower macro metrics on rare classes (e.g., Chapter IX blocks).<br>- No external validation beyond Portuguese data.<br>- Focused on short texts; not suited for long clinical documents. | 2, 3 |
| 21 | - Proposed BW_att, a new model combining BERT, Word2Vec, and label attention, tailored for CHD coding.<br>- Outperformed existing baselines (e.g., LAAT, CAML) in most tasks.<br>- Achieved high interpretability by identifying key predictive tokens in clinical text.<br>- Demonstrated strong performance even on long, complex discharge summaries.<br>- Conducted ablation studies confirming the value of their truncation method. | - Evaluated only on two datasets (Fuwai, MIMIC-III); portability across hospitals not tested.<br>- Focused on frequent CHD codes; rare codes were excluded to avoid overfitting.<br>- Did not explore generalization across different coding systems or specialties.<br>- Plans for future work include expanding to rare codes using GNNs or hybrid rule-based methods. | 2 |
| 25 | - Outperformed baseline BERT in all settings, with major gains in Precision (e.g., +44.6 pts in Spanish, >100% in Swedish).<br>- Introduced PlaBERT, a novel per-label attention mechanism enabling explainable predictions.<br>- Achieved high performance on multilingual EHRs, including low-resource languages like Swedish and Spanish.<br>- Developed new metrics (Alignment Precision/Recall) to assess EHR coding quality.<br>- Released open-source implementation compatible with BERT, RoBERTa, XLNet, XLM, and DistilBERT. | - Performance limited by BERT's maximum sequence length and diluted attention for broad ICD codes.<br>- Original Swedish dataset had poor alignment between codes and text, affecting performance.<br>- No real-world clinical validation with physicians or coding professionals.<br>- No support for full ICD-10 coverage beyond gastrointestinal (K-codes). | 2, 3 |
| 28 | - Demonstrated ChatGPT (GPT-3.5) can generate at least one correct ICD-10 code in 70% of retina clinical encounters without feedback.<br>- Achieved 59% completely correct coding rate.<br>- Could handle laterality-specific coding (right vs. left eye) in most cases.<br>- Highlighted potential for reducing physician coding burden and improving clinic workflow.<br>- Identified areas where LLMs may assist, not replace, clinician final review. | - ChatGPT hallucinated nonexistent ICD-10 codes in some cases.<br>- Misassigned laterality (right/left eye) codes 6.6% of the time.<br>- Performance deteriorated without resetting the chat between cases (sequential memory effect).<br>- No fine-tuning or real-world billing validation performed.<br>- Only mockup data used — no real patient records or external hospital validation.<br>- Model based on GPT-3.5, not updated with latest ICD-10 revisions. | 2 |
| 24 | - Successfully applied transformer model to 5.3M death certificates.<br>- Achieved high shortlist-level F1-score (0.94).<br>- Enabled full national coding for 2018–2019. | - Lower recall for musculoskeletal and blood diseases.<br>- Rare codes showed higher error rates.<br>- Lack of interpretability analysis. | 2,3 |
| 27 | - High accuracy (Q1: 94.4%) and sensitivity (98.2%) in detecting GI bleeding. | - Single clinical task and center (GI bleeding, MIMIC-III). | 2 |
| | - Outperformed ICD-9 in all tasks.<br>- Comparable to human reviewers.<br>- Fast and cost-efficient (12.7 min, \$0.33/chart).<br>- Ensured HIPAA-compliant data privacy. | - Used ICD-9, not ICD-10.<br>- Errors in complex or ambiguous cases.<br>- Some charts excluded due to token limits.<br>- No comparison with other LLMs. | |
| 35 | - First to assess ChatGPT on nephrology ICD-10 coding<br>- GPT-4 reached 99% accuracy across both rounds<br>- Realistic pre-visit case scenarios<br>- Simple prompt design fits clinical workflow<br>- Potential to ease documentation burden | - Used simulated cases, not real EMRs<br>- No comparison to human or rule-based coding<br>- Tested only single-code assignments<br>- Limited to nephrology; no broader validation | 2 |
| 37 | – Introduces a novel summarization-based ICD coding model (SuM) combining a pointer network with text matching. – Achieves high performance on both TOP50 (AUC: 0.9548) and FULL (AUC: 0.9124) datasets. – Outperforms XR-LAT and BM25 baselines in all metrics. – Ablation shows end-to-end structure and pointer network improve accuracy. | – Evaluated only on Chinese ICD-10 neoplasm codes (C00–C97) from a single oncology department. – No testing on other ICD chapters or external datasets. – No multilingual evaluation or interoperability testing.<br>– Does not assess prediction time or integration into clinical workflows. | 2,3 |
| 30 | – Demonstrated that using a hierarchical, multi-task approach improves consistency and accuracy of ICD predictions across Full, Main, and Chapter levels.<br>– Validated the use of in-house, in-domain Spanish EHRs to enrich language models in resource-limited settings.<br>– Released reproducible software for multi-task classification compatible with various transformer models. | – Limited to Spanish EHRs from a single institution, raising concerns about generalizability.<br>– Performance at the most detailed ICD level (Full) remains significantly lower than at Chapter level.<br>– Lack of external validation or multilingual testing limits the broader applicability of findings. | 3 |

## Data Availability

All data produced in the present work are contained in the manuscript

