## Supplemental Tables for "Automatic ICD coding using LLMs: a systematic review"

**Supplementary table S1: Search strategies used to identify studies on large language models for ICD coding.**

| **Database** | **Search strategy** |
| --- | --- |
| PubMed | ((((((OpenAI) OR (ChatGPT)) OR (LLM)) OR ("large language model")) OR ("Microsoft Bing")) OR ("Google Gemini")) OR (transformer) OR (BERT)) AND ((ICD) OR ("International Classification of Diseases")) |
| Embase | ('openai' OR 'chatgpt' OR 'llm' OR 'large language model' OR 'microsoft bing' OR 'google gemini' OR 'transformer' OR 'bert') AND ('international classification of diseases' OR 'icd') |
| Google Scolar | ("OpenAI" OR "ChatGPT" OR LLM OR "large language model" OR "Microsoft Bing" OR "Google Gemini" OR transformer OR BERT) AND (ICD OR "International Classification of Diseases") |

**Supplementary Table S2: Inclusion and Exclusion Criteria for Studies on LLM-Based ICD Coding**

| **Criterion** | **Inclusion** | **Exclusion** |
| --- | --- | --- |
| Study design | Original research, peer-reviewed articles | Reviews, editorials, commentaries, non-peer-reviewed publications, conference abstracts |
| Model type | Studies applying large language models (BERT, GPT, Google Gemini, Microsoft Bing) for ICD coding | Manual coding only; non-LLM automated methods (e.g. rule-based, SVM, random forest); other coding systems (CPT, SNOMED) |
| Data source | Clinical or administrative data (EHR notes, discharge summaries, other medical documentation) | Non-clinical applications; datasets unrelated to ICD coding |
| Comparators | Human/manual coding or legacy automated systems (if reported) | n/a (absence of a comparator does not exclude) |
| Outcomes | Reports ≥ 1 performance metric (accuracy, precision, recall, F1 score, coding time, user satisfaction) | No quantitative or qualitative performance evaluation |
| Language | English-language studies | Non-English publications |
| Date range | Studies published up to August 2024 | Studies published after August 2024 (per search end |

**Examples of codes and prompts:**

1. **Can GPT-3.5 generate and code discharge summaries?**

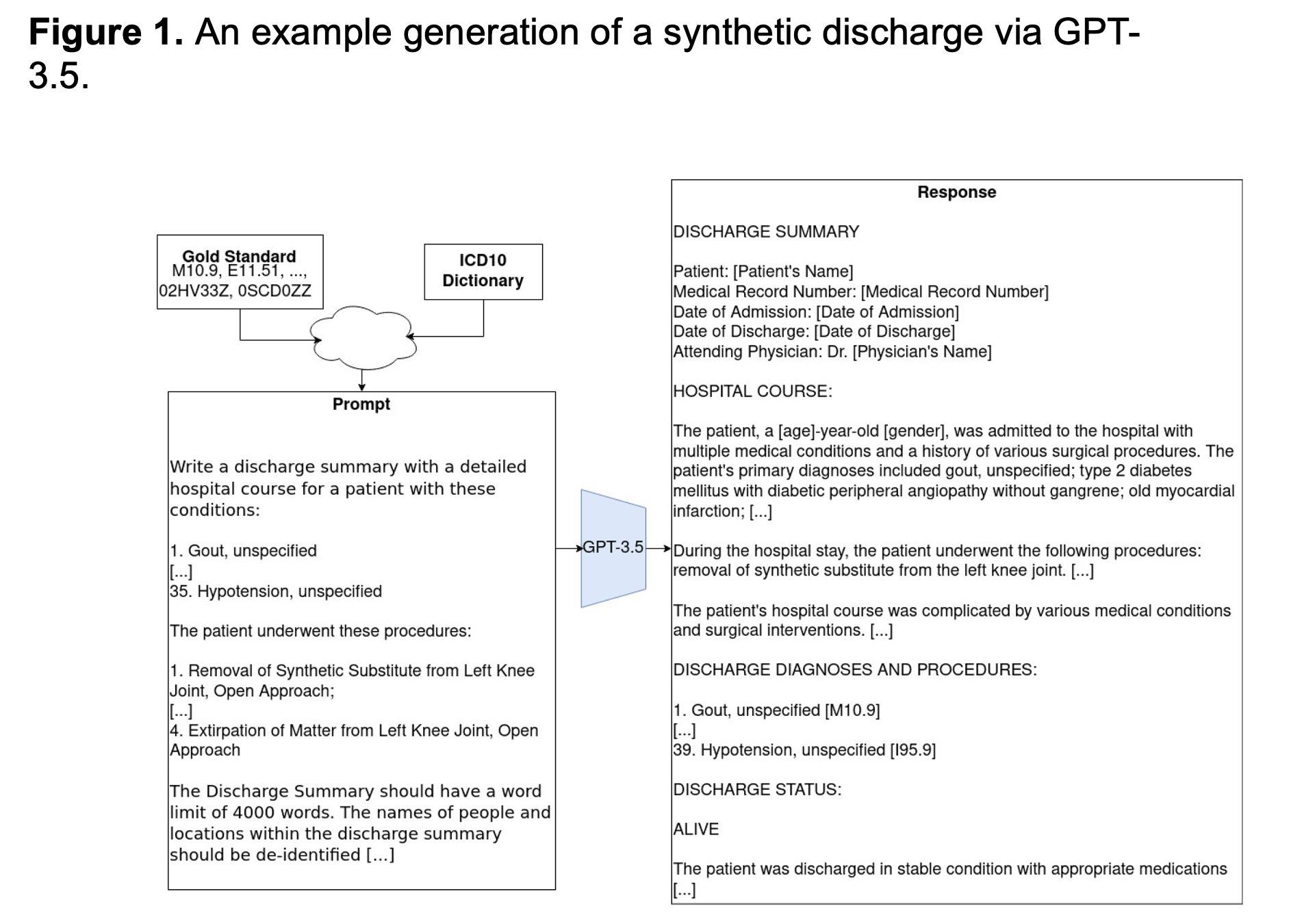

2. **Aiding ICD-10 Encoding of Clinical Health Records Using Improved Text Cosine Similarity and PLM-ICD**

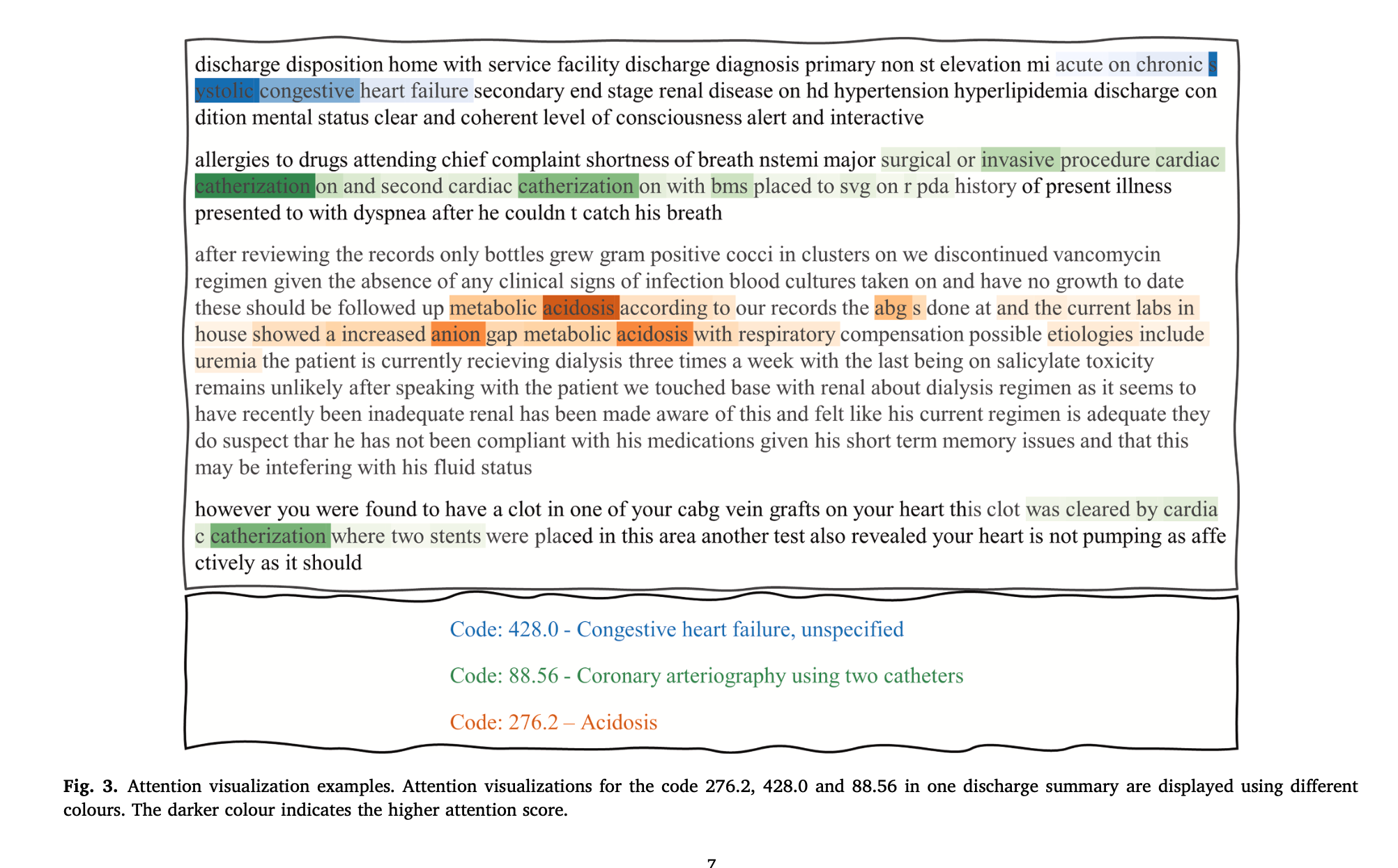
